# Platelet dysfunction in immune thrombocytopenia: finding clinical subsets with platelet phenotypes

**DOI:** 10.1101/2025.07.15.25331619

**Authors:** Sidra A. Ali, Sarah M. Hicks, Lucy A. Coupland, Simone A. Brysland, Vijay Bhoopalan, Yee Lin Thong, Robert K. Andrews, Elizabeth E. Gardiner, Philip Y-I. Choi

**Author notes:** These authors contributed equally (Sidra A. Ali, Sarah M. Hicks). Corresponding Authors: Associate Professor Philip Choi.Professor Elizabeth Gardiner.

## Abstract

Patients with immune thrombocytopenia (ITP) remain a challenge to diagnose, manage, and predict bleeding risk. A comprehensive assessment of platelet function may aid clinical management. This study assessed platelet parameters to predict bleeding in ITP. Blood from 103 clinically-annotated cases with isolated thrombocytopenia or 123 healthy donors was evaluated. In the ITP cohort, 75/110 encounters (68%) had platelet counts below 50 x 10⁹/L. Platelet surface proteins, reticulated platelets, and activation were quantified using flow cytometry. Soluble receptor fragments, citrullinated histone-DNA (CitH3-DNA) complexes, and thrombopoietin (TPO) were quantified by ELISA. Whole blood clotting and platelet contribution to clot formation were evaluated using viscoelastography. Elevated levels of glycoprotein (GP) VI (p=0.0012), Trem-like transcript-1 (TLT-1) (p=0.0248), platelet-bound immunoglobulin (Ig) G (p<0.0029), CitH3-DNA complexes (p=0.0022), TPO (p<0.0001), and reduced platelet contribution to clot formation (p<0.0001), were observed in primary ITP patients with bleeding and bruising symptoms. A multivariable analysis revealed that measuring platelet indices strengthened a predictive bleeding model over platelet count alone (78.1% vs. 70.48%). Symptomatic ITP patients have measurable platelet dysfunction and quantifiable differences on platelet surface, increased evidence of NETosis and elevated TPO levels. Identifying biomarkers for ITP outcomes can define subsets of disease with clinical relevance.

Visual abstract **Key biomarkers and assays evaluated in this study**. The top left panel depicts flow cytometry-based approaches performed in whole blood samples for studying platelet surface proteins, including integrins, glycoproteins, and activation markers, as well as the assessment of pathways regulating integrin αIIbβ3 activation. The right panel highlights ELISA-based detection of soluble fragments (sTLT-1, sGPVI) released from activated platelets, along with serum TPO and NET formation in plasma and serum samples. The bottom panel illustrates ROTEM analysis in normal clot formation, ITP patients with thrombocytopenia and its ability to detect platelet dysfunction.

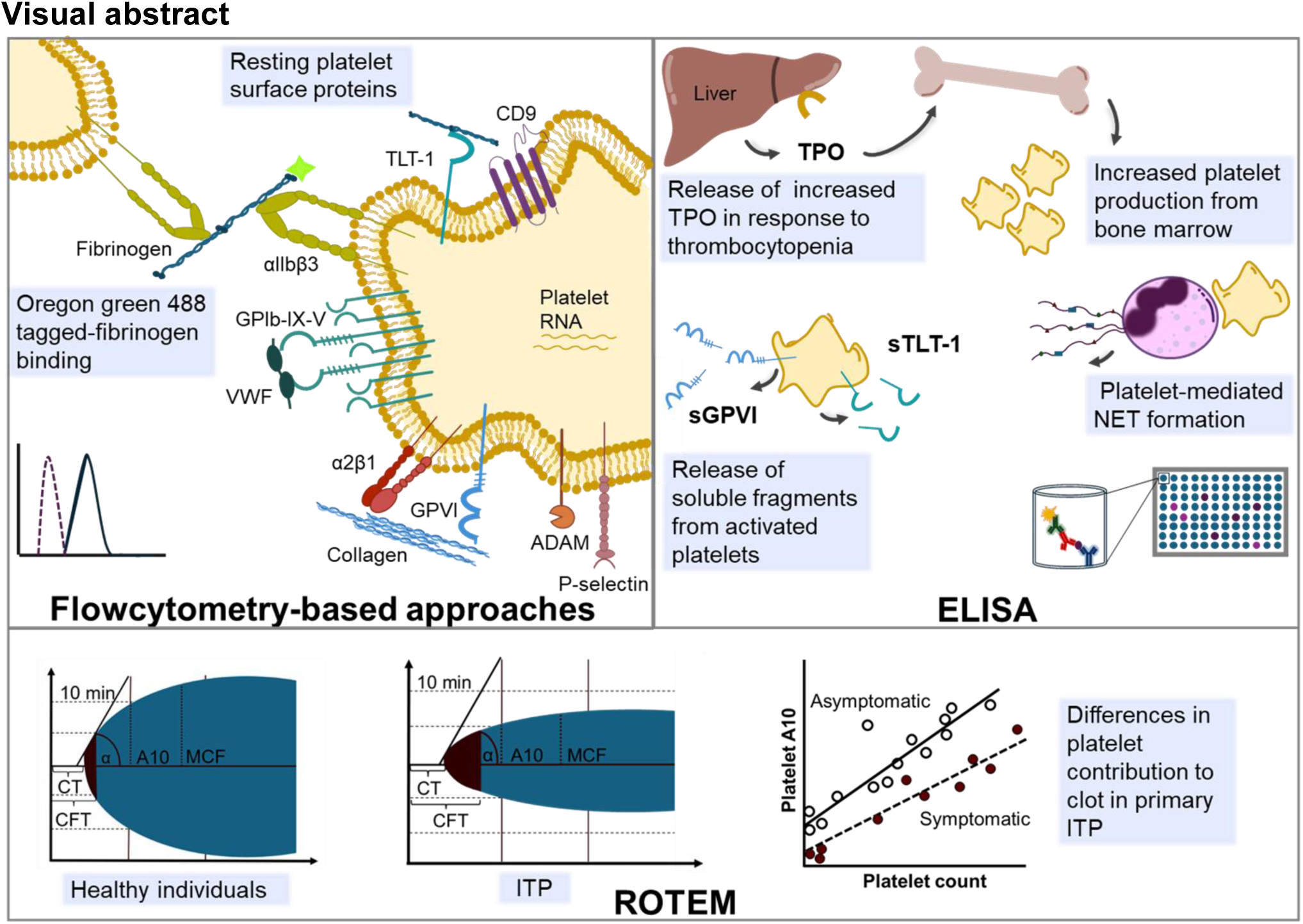

## Introduction

Immune thrombocytopenia (ITP) is a rare disease, with an estimated incidence in adults ranging from 1.6 to 3.9 cases per 100,000 per year.^1^ Insights into the pathogenic mechanisms underlying ITP and the advent of therapies, including thrombopoietin receptor agonists (TPO-RA) and rituximab, have changed the landscape of ITP management. Still, a challenging minority of patients remain refractory to multiple lines of treatment and develop chronic forms of thrombocytopenia, repeated bleeding episodes, poor health-related quality of life, and high mortality.^2^ ITP is a broad and imprecise diagnostic classification used to describe several molecularly distinct disorders^3^ that may include varying contribution by platelet specific autoantibodies, loss of regulation of immune function, and acquired megakaryocyte maturation abnormalities.^4,5^

Platelets maintain blood volume and aid innate immune responses and wound repair.^6^ In ITP, key platelet receptors including glycoprotein (GP) Ib-IX-V complex, GPV, GPVI, and integrins αIIbβ3 and α2β1 are targeted by antiplatelet autoantibodies^7^ which can mediate platelet activation via Fc-dependent and -independent means.^8^ Major challenges in ITP include a paucity of definitive diagnostic tools and a lack of clinical acuity regarding the ideal therapeutic option for each patient. As platelets are targeted in ITP, assessing their function makes sense but is challenging as samples need to be collected and rapidly analysed ideally using assays that are insensitive to platelet numbers.

Rapid, point-of-care testing of platelet function also provides significant benefit to determine bleeding risk, as shown in a pilot study of ITP cases with comparable thrombocytopenia.^9^ Non-time sensitive measures of platelet activation and function are a clinical unmet need. Flow cytometry is an appealing alternative, as it is largely unaffected by the platelet count. Further, platelet binding by anti-platelet autoantibodies which is central to ITP pathogenesis,^1,10–12^ can cause rapid and irreversible proteolytic release of receptor ectodomains such as soluble GPVI (sGPVI) and soluble triggering receptor expressed on myeloid cells-like (Trem)-like transcript-1 (sTLT-1).^13^ Measurement of sGPVI and sTLT-1 reflect platelet activation and degranulation. Activated platelets also promote the formation of neutrophil extracellular traps (NETs), which are implicated in immune-mediated cellular destruction and correlate with clinical severity.^14^ These biomarkers were selected for their relevance to known ITP mechanisms, including immune-mediated platelet activation, receptor shedding, and platelet–immune cell interactions.

Our objective here was to combine flow cytometric evaluation of platelet quality and function, along with viscoelastography and measurement of soluble plasma or serum proteins in blood drawn from ITP patients, to develop a platelet signature that could aid clinical diagnosis, predict clinical course, and ultimately help stratify patients with ITP for appropriate therapy. With the range of emerging targetable aspects of ITP,^15–18^ diagnostic tools to aid the logical selection of therapy become increasingly imperative.

## Methods

### Whole blood collection and clinical annotations

The National Platelet Research and Referral Centre (NPRC) is a specialist centre for platelet evaluation within the John Curtin School of Medical Research and Canberra Health Services. The NPRC hosts a biobank of plasma samples and an annotated clinical database that documents bleeding episodes, disease duration and treatment responses (Table S1). Non-consecutive patients were referred for NPRC biobanking with disorders of platelet number (< 100 x 10^9^/L) or suspected platelet dysfunction. This study received Human Research Ethics Committee approval from ANU (2022/372 and 2017/924) and ACT Health (ETHLR.18.058). All patients and healthy donors (HDs) provided informed consent prior to participation, and the study was conducted as per the Declaration of Helsinki under the NPRC protocol at The Canberra Hospital.

Patients with WHO bleeding (≥ grade 1) or ITP-BAT score ≥ 1,^19^ were pooled and compared to asymptomatic patients. Patients receiving steroids and/or intravenous immunoglobulin (IVIg) were classified as undergoing first-line therapy, while those receiving any other treatments were grouped under second-line therapy. This included TPO-RA, rituximab, mycophenolate mofetil, dapsone, cyclosporine, danazol, vincristine, azathioprine, oseltamivir, ianalumumab, and efgartigimod or splenectomy.

### Flow cytometry analysis

Details can be found in Supplementary Methods. Reticulated platelets were quantified using thiazole orange (TO). To quantify platelet surface proteins, trisodium citrate (TSC)-anticoagulated whole blood (WB) was incubated with fluorescently-conjugated antibodies against platelet-specific proteins and antibody-bound platelet events were acquired (Figure S1). To assess platelet αIIbβ3 activation (Figure S2A for gating strategy), WB was stimulated with platelet agonists in the presence of Oregon-Green-tagged fibrinogen (OG-Fg). FlowJo™ software (v10.8 BD Life Sciences) was used for analysis.

### Viscoelastography with ROTEM

All samples also underwent ROTEM analysis within four hours of venepuncture. To gain an indication of the platelet contribution to clot formation, platelet A10 was calculated by subtracting FibTEM A10 from EXTEM A10. FibTEM uses cytochalasin D, an actin polymerisation inhibitor, to block platelet cytoskeletal changes, resulting in a fibrin-dominant clot. In contrast, EXTEM reflects both platelet and fibrin contributions.

Detailed descriptions of all other experimental methods are provided in the Supplementary Methods.

### Statistical analysis

Details can be found in Supplementary Methods. Box plots depict median, interquartile ranges, and extremes.

## Results

### NPRC patient cohort overview

The NPRC investigates platelet phenotypes in patients with thrombocytopenia using research-based assays. Between 1^st^ October 2016 and 1^st^ April 2024, 103 patients were recruited, and blood samples and clinical data were collected. Cohort characteristics (Figure 1) show 68% (70/103) of patients were diagnosed with primary ITP (pITP), 10% (11/103) with secondary ITP (sITP) with a known underlying aetiology, and 21% (22/103) with platelet counts below 100 x 10^9^/L due to causes other than ITP (control thrombocytopenic group; Table S2).

**Figure 1.**
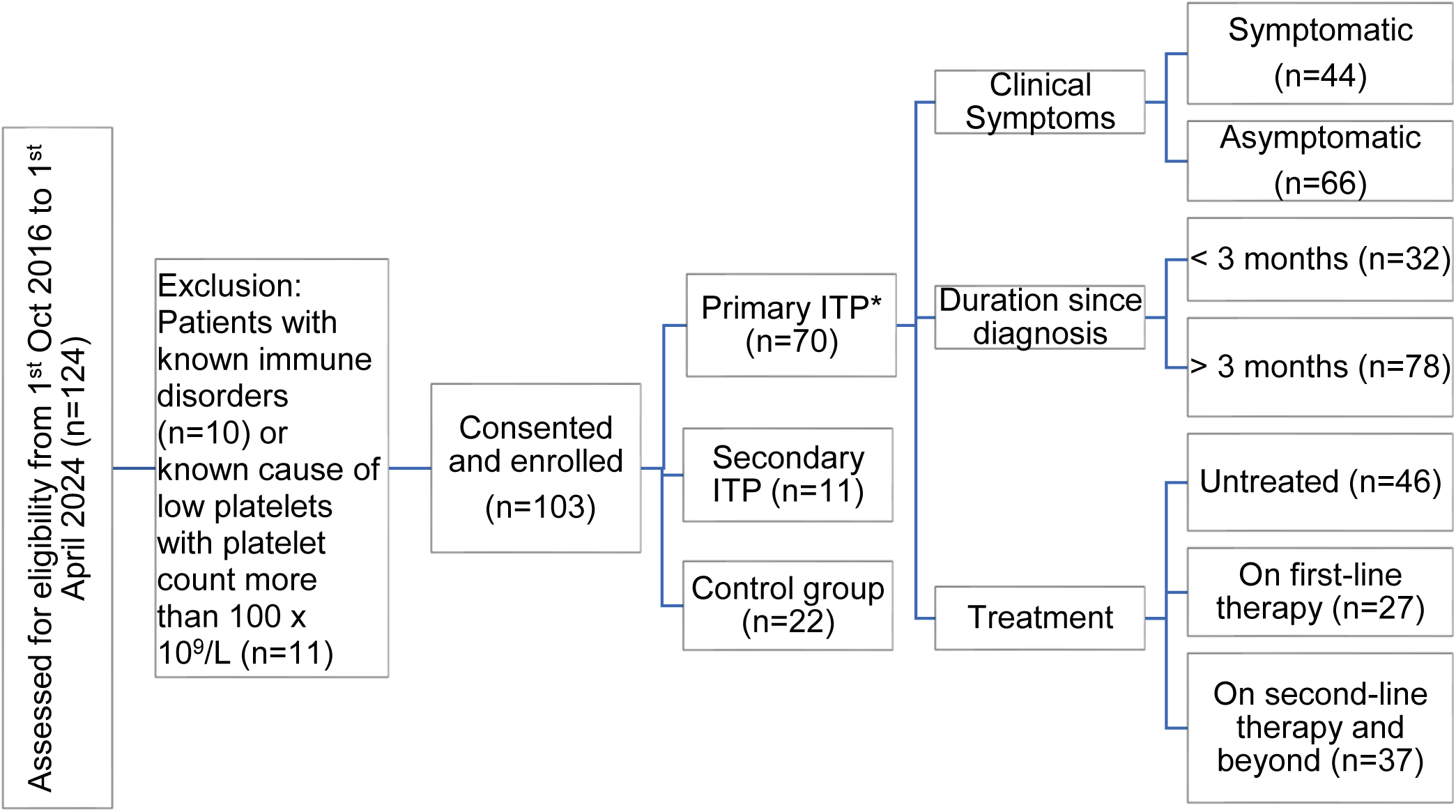
Patient recruitment flow chart and demographics. ITP=Immune thrombocytopenia, control group=patients with platelet count below 100 x 10^9^/L due to causes other than ITP, MPV=mean platelet volume. Patients receiving steroids and/or IVIg at the time of blood collection were classified as undergoing first-line therapy, while the remaining patients were receiving second-line therapy and beyond. *110 encounters from primary ITP patients, involving 70 individual patients, with repeated sample collections (n=21) at different time points, including during remission.

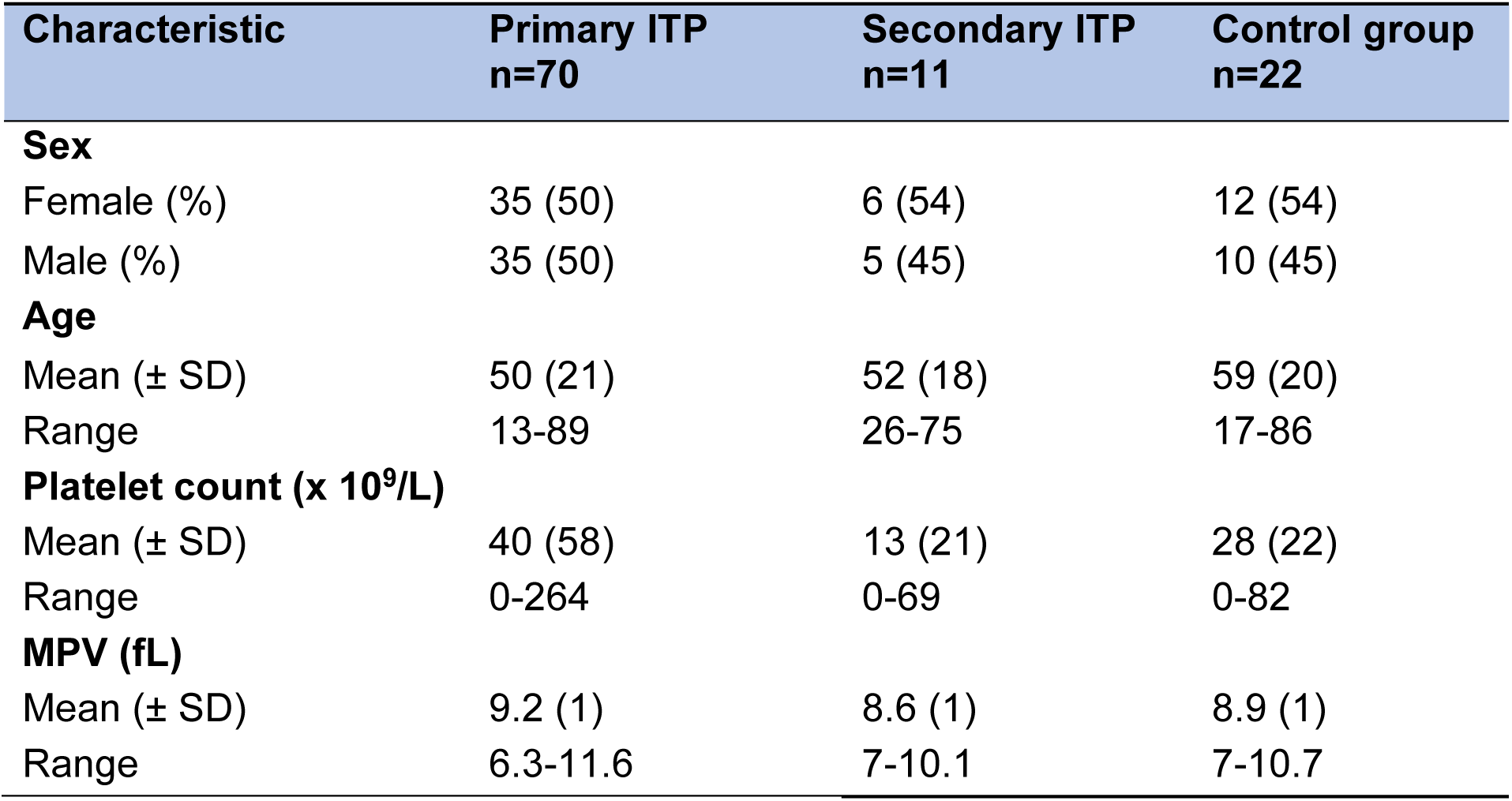

All patient groups (pITP, sITP, and the control group) had significantly lower platelet counts (p<0.0001 for all, Figure 2A) and increased MPV (p<0.0001, 0.0023, and p<0.0001, respectively, Figure 2B) compared to HDs. Within pITP cohort, platelet counts were significantly lower in symptomatic pITP (p<0.0001, Figure 3A), newly diagnosed (p<0.0489, Figure S3A), and in those receiving steroids and/or IVIg (p<0.0021, Figure 4A). However, many patients with low platelet counts did not experience any bleeding or bruising symptoms, consistent with hypotheses that patients experiencing bleeding tendencies had reduced platelet function.^20^ Differences remained non-significant even after adjustment for platelet numbers (data not shown). Newly released RNA-enriched (TO^bright^) platelets showed no difference between HDs, ITP and control groups (Figure 2C) or across bleeding symptoms (Figure 3C), stage (Figure S3C), or first-line treatments (Figure 4C). However, pITP patients showed a mild but significant correlation (r=0.435, p=0.030) between TO^bright^ events and surface GPIbα levels (Figure S4), aligning with higher GPIbα on younger platelets.^21^

**Figure 2:**
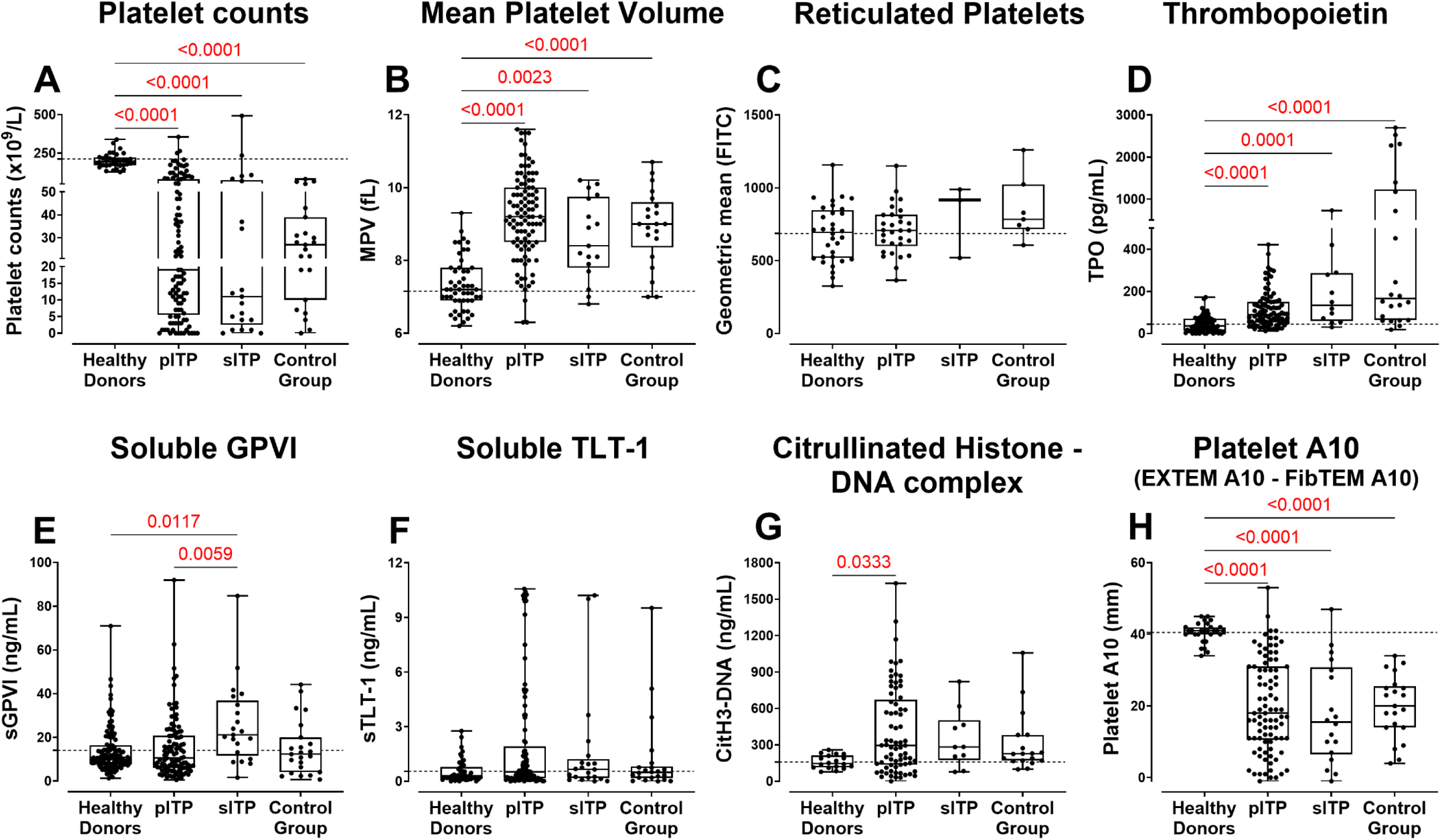
Platelet and plasma parameters in healthy donors, ITP and control group. **A.** Platelet counts and **B.** MPV were measured using an automated haematology analyser in TSC-anticoagulated WB. **C.** Reticulated platelets were quantified by TO staining in flow cytometry. Levels of **D.** thrombopoietin (TPO), **E.** soluble (s) GPVI, **F.** sTLT-1 and **G.** citrullinated histone-DNA (CitH3-DNA) complex were measured by ELISA. **H.** Platelet A10 was calculated by subtracting the FibTEM A10 measurement from the EXTEM A10 in ROTEM in healthy donors (n=18-123), pITP patients (n=30-105), sITP (n=3-12) and control group (n=7-23). Only significant p-value (p<0.05) highlighted in red are displayed. Data includes repeat measurements from the same subjects collected at different time points. The dotted horizontal line represents the healthy donor mean measured alongside these patients. pITP=primary ITP, sITP=secondary ITP.

**Figure 3.**
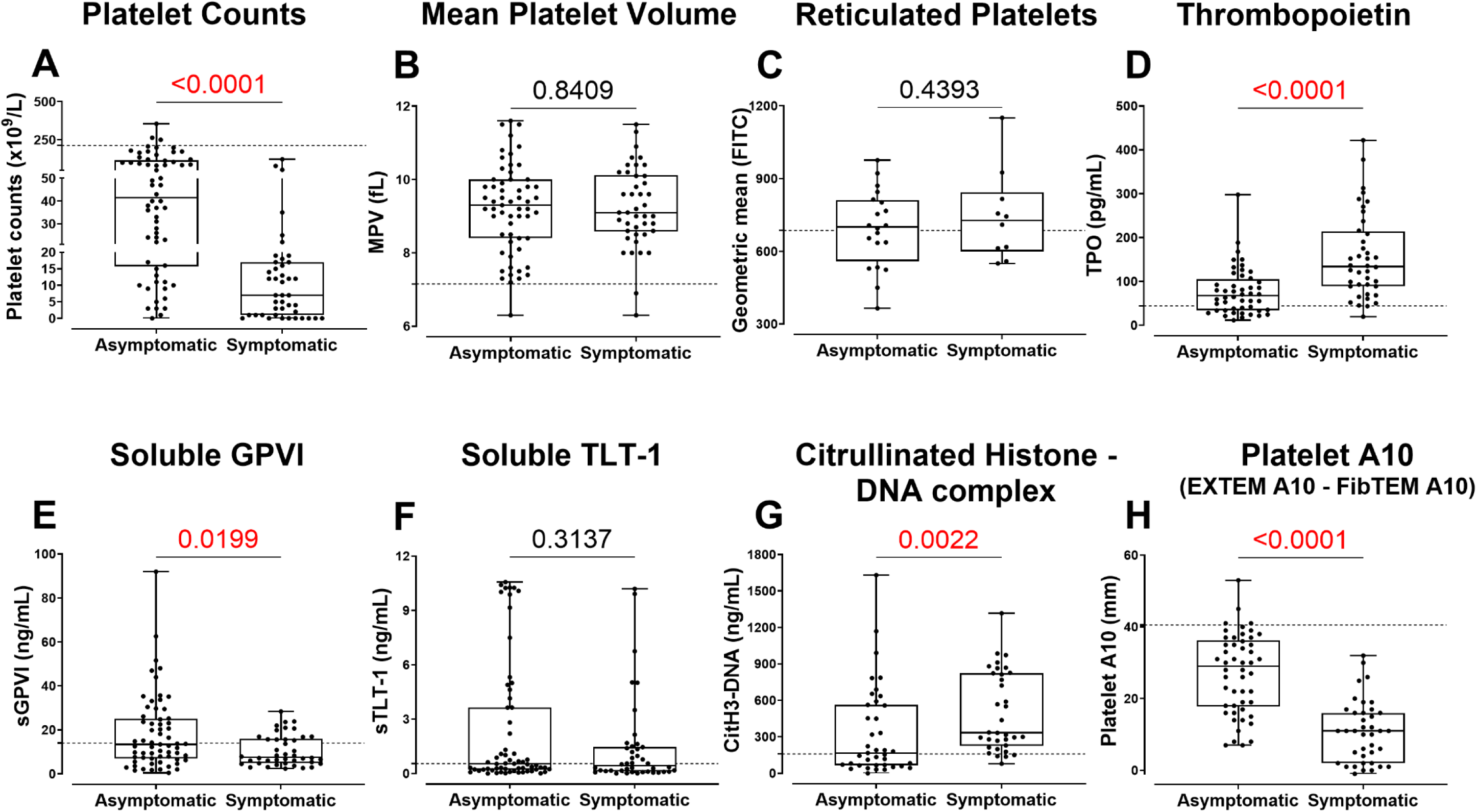
Platelet and plasma parameters in primary ITP patients with or without clinical symptoms of bleeding or bruising. **A.** Platelet counts and **B.** MPV were measured using an automated haematology analyser in TSC-anticoagulated WB. **C.** Reticulated platelets were quantified by TO staining in flow cytometry. Levels of **D.** thrombopoietin (TPO), **E.** soluble (s) GPVI, **F.** sTLT-1 and **G.** citrullinated histone-DNA (CitH3-DNA) complex were measured by ELISA. **H.** Platelet A10 was calculated by subtracting the FibTEM A10 measurement from the EXTEM A10 in ROTEM. The primary ITP patient cohort (n=10–63) includes repeat measurements from the same subjects collected at different time points. An unpaired t-test or Mann-Whitney test was performed depending on the distribution of the data. Significant p-values (p<0.05) are highlighted in red. The cutoff for HD levels is indicated by dotted horizontal lines.

**Figure 4.**
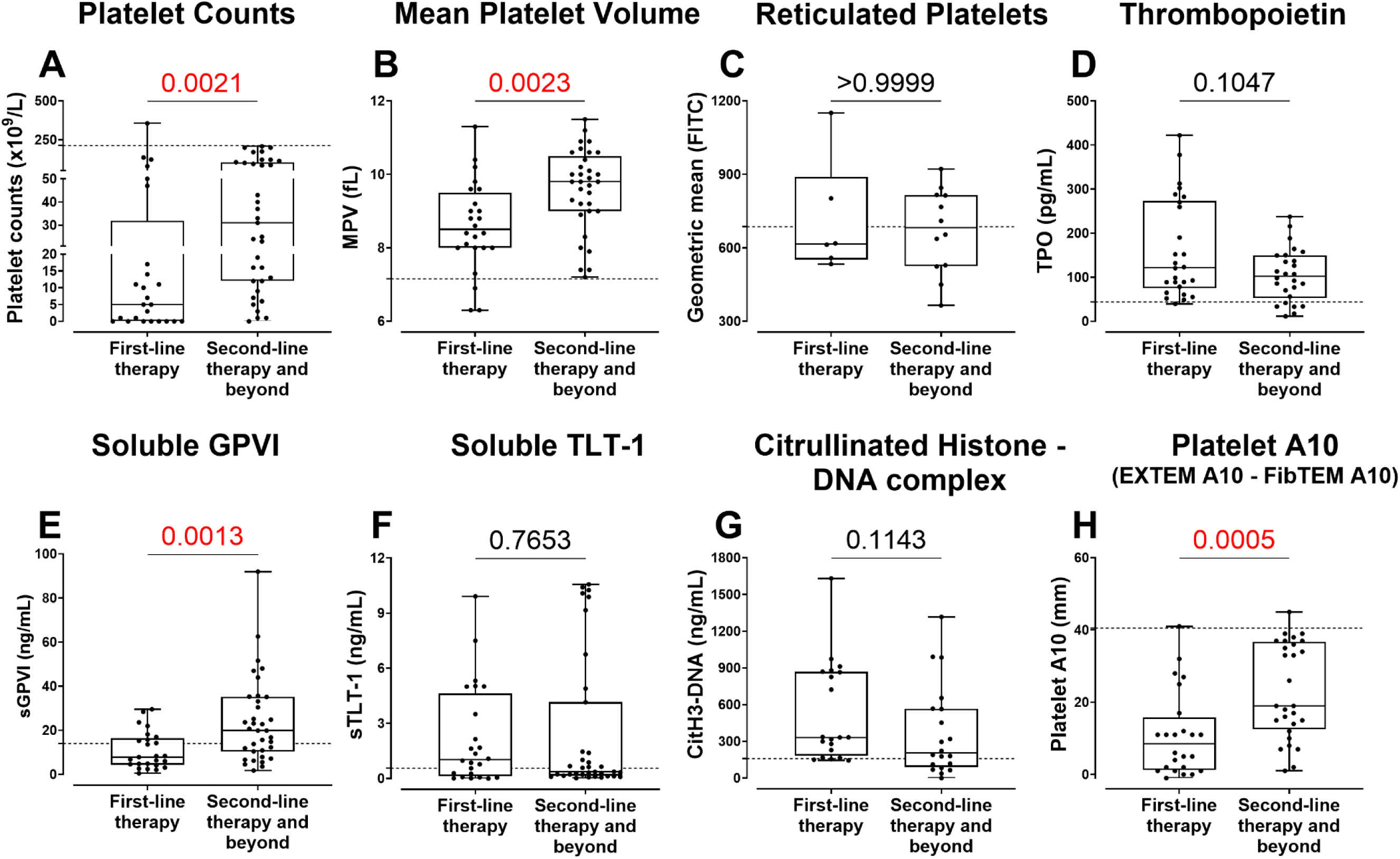
Platelet and plasma parameters in primary ITP patients stratified for therapy. Patients receiving first-line therapy (steroids and/or IVIg) were compared to those receiving second-line therapy and beyond (which include first line therapy plus treatments with TPO-RA, rituximab, mycophenolate mofetil, dapsone, cyclosporine, danazol, vincristine, azathioprine, oseltamivir, ianalumumab, and efgartigimod as well as splenectomy). **A.** Platelet counts and **B.** MPV were measured using an automated haematology analyser in TSC-anticoagulated WB. **C.** Reticulated platelets were quantified by flow cytometry. Levels of **D.** TPO, **E.** sGPVI, **F.** sTLT-1 and **G.** CitH3-DNA complexes were measured by ELISA. **H.** Platelet A10 was calculated by subtracting the FibTEM A10 measurement from the EXTEM A10 in ROTEM. The primary ITP patient cohort (n=6-35) includes repeat measurements from the same subjects collected at different time points. An unpaired t-test or Mann-Whitney test was performed depending on the distribution of the data. Significant p-values (p<0.05) are highlighted in red. The cutoff from HD levels is indicated by a dotted horizontal line on the respective graphs.

### Serum TPO levels were elevated in symptomatic thrombocytopenic patients

The pITP, sITP and control group had high serum TPO concentrations compared to HDs (p<0.0001 for all, Figure 2D). In the pITP group, TPO levels showed a negative correlation with platelet count (r=-0.435, p<0.0001, Figure S5A) and were significantly elevated in patients with platelet counts below 20 x 10⁹/L (Figure S5B) as well as in those experiencing bleeding or bruising at the time of collection (p<0.0001; Figure 3D). No significant difference in TPO levels was observed between pITP patients receiving TPO receptor agonists and those on other treatments (Figure S5C).

### Plasma sGPVI and sTLT-1 in pITP

Elevated sGPVI levels were observed in asymptomatic pITP (p=0.0199, Figure 3E) and patients receiving second-line line therapy and beyond (p=0.0013, Figure 4E). No correlation was observed between sGPVI or sTLT-1 with platelet counts (Figures S6B and S6C) or their respective surface receptors (data not shown), suggesting that proteolysis of these receptors was not constitutive and likely to be linked with platelet activation status. Consistent with this, a mild but significant correlation was found between sGPVI and sTLT-1 levels (r=0.302, p=0.002, Figure S6A) in pITP samples. To further investigate the influence of platelet count and clinical features, both soluble markers were analysed across subgroups stratified by platelets and clinical parameters; however, no significant differences were observed (data not shown). No differences in the soluble markers were observed in pITP when compared to HDs or control group (Figures 2E and 2F), however sITP showed elevated sGPVI when compared with pITP (p=0.0059, Figure 2E) and HDs (p=0.0117, Figure 2E).

### Surface GPVI and TLT-1 levels were elevated in ITP patients with bleeding

Most of the platelet surface proteins remained unaltered in pITP patients when compared within subgroups; however, GPVI, TLT-1 and antihuman antibody binding was significantly elevated in patients exhibiting clinical symptoms (p=0.0012, p=0.0248 and p=0.0029 respectively, Table 1). When compared with HDs, pITP patients showed increased expression of platelet activation markers P-selectin and TLT-1 (p=0.0084 and p=0.0136, respectively, Table S3) and antihuman antibody binding (p=0.0005, Table S3) despite the heterogeneity of cohort.

**Table 1:** Platelet surface protein expression in primary ITP patients based on clinical presentation, disease chronicity and treatment.

| Primary ITP* | Surface proteins changes in geometric mean (P-value) |  |  |  |  |  |  |  |  |
| --- | --- | --- | --- | --- | --- | --- | --- | --- | --- |
| | GPVI<br>n=7-41 | GPIbα<br>n=7-41 | Integrin<br>αIIb<br>n=7-41 | Integrin<br>α2<br>n=6-27 | CD9<br>n=6-29 | ADAM10<br>n=6-28 | P-selectin<br>n=6-28 | TLT-1<br>n=7-38 | Anti-human<br>IgG<br>n=7-36 |
| **Symptoms** |  |  |  |  |  |  |  |  |  |
| (Symptomatic vs asymptomatic) |  |  |  |  |  |  |  |  |  |
| P-value | 0.0012 | 0.9522 | 0.2066 | 0.5102 | 0.7733 | 0.5019 | 0.3460 | 0.0248 | 0.0029 |
| Change | Increased | No change | No change | No change | No change | No change | No change | Increased | Increased |
| **Stage** |  |  |  |  |  |  |  |  |  |
| (< 3months vs >3 months) |  |  |  |  |  |  |  |  |  |
| P-value | 0.8982 | 0.7227 | 0.1216 | 0.2686 | 0.8456 | 0.3635 | 0.8658 | 0.2893 | 0.3744 |
| Change | No change | No change | No change | No change | No change | No change | No change | No change | No change |
| **On first-line treatment†** |  |  |  |  |  |  |  |  |  |
| (Yes vs. No) |  |  |  |  |  |  |  |  |  |
| P-value | 0.0654 | 0.9925 | 0.0541 | 0.2483 | 0.7378 | 0.8236 | 0.1505 | 0.0717 | 0.3760 |
| Change | No change | No change | No change | No change | No change | No change | No change | No change | No change |

\*Includes repeat measurements from the same patient on different days including during recovery. An unpaired t-test or Mann-Whitney test was performed depending on distribution of the data. P-values highlighted in red are significant (p<0.05). ITP=immune thrombocytopenia, GP=glycoprotein, CD=cluster of differentiation, ADAM=a disintegrin and metalloproteinase, TLT-1=TREM like transcript-1, IgG=immunoglobulin G.†Patients receiving steroids and/or IVIg at the time of blood collection were classified as undergoing first-line therapy, while the remaining patients were receiving second-line or subsequent therapies.

### ITP patients demonstrated normal function of pathways controlling integrin αIIbβ3 **activation**

Platelets from pITP patients responded effectively to low and high doses of cross-linked collagen-related peptide (CRP-xL) and adenosine diphosphate (ADP) in OG-Fg activation assay (Figure S2). There were no differences observed between HDs and pITP patients or within ITP subgroups (data not shown).

### CitH3-DNA complex levels were elevated in symptomatic ITP patients

Increased CitH3-DNA complex levels were noted in bleeding or bruising pITP patients (p<0.0022, Figure 3G). The effect of therapy on CitH3-DNA complex levels was investigated and although the difference was not statistically significant (p=0.1143, Figure 4G), higher levels were observed in ITP patients undergoing first-line treatment. No significant correlation was observed between CitH3-DNA complex levels and platelet count (Figure S7A-B), indicating that elevated CitH3-DNA levels in symptomatic patients are independent of platelet mass.

### ROTEM parameters can stratify ITP patients at greater risk of bleeding

Clot formation time (CFT) was significantly prolonged in pITP patients compared to HDs in EXTEM (p=0.0099) and INTEM (p=0.0032, Figure 5A). pITP samples also showed reduced clot amplitude at 10 minutes (A10), and maximum clot firmness (MCF) EXTEM, INTEM, and NATEM (data not shown).

**Figure 5.**
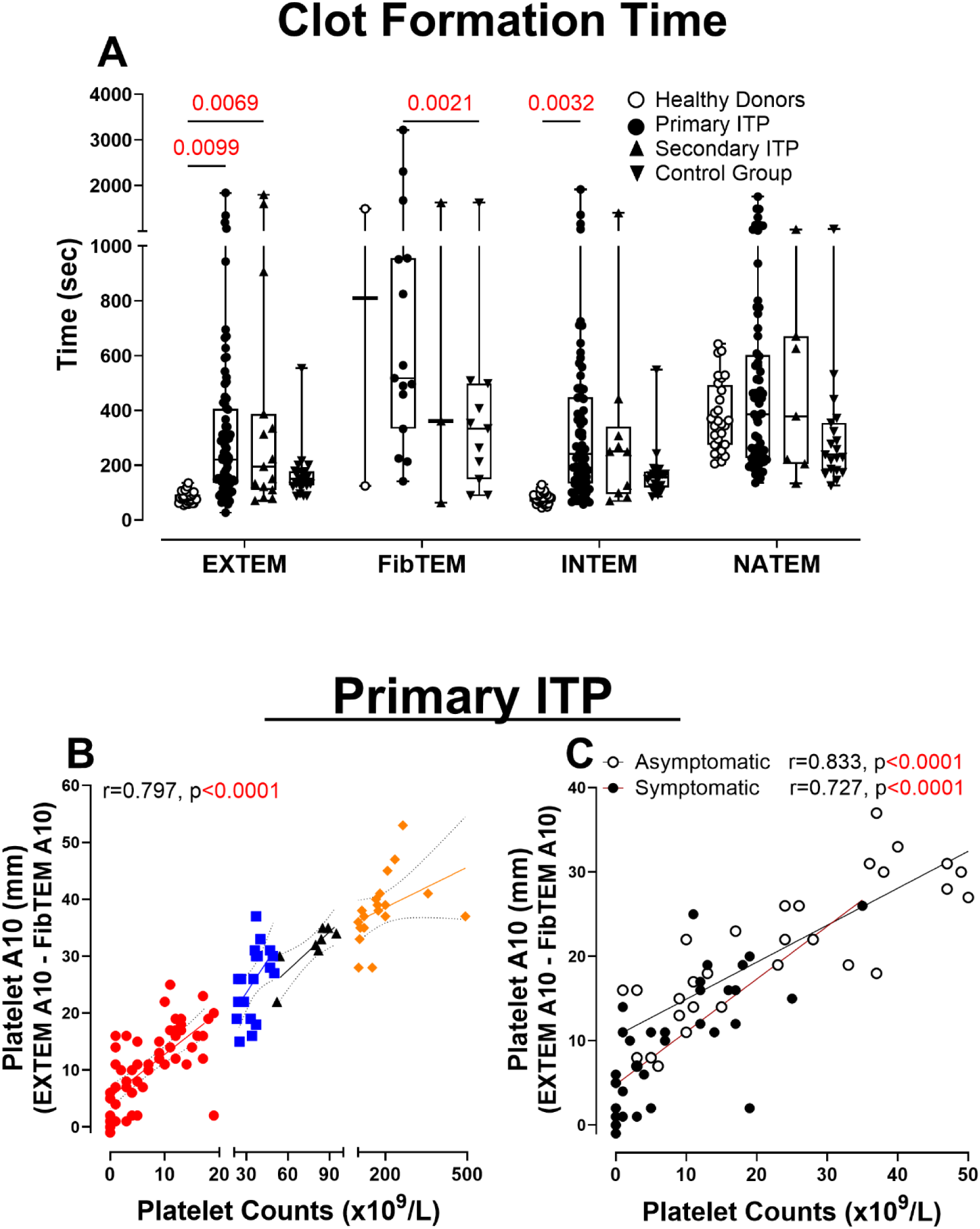
Platelet contribution to clot formation in primary ITP patients. **A.** Clot formation time - time from initiation of clotting until a clot firmness of 20 mm is detected. Platelet A10 was calculated by subtracting the FibTEM A10 measurement from the EXTEM A10. **B.** Correlation between platelet A10 and platelet counts in ITP patients with platelet counts below 20 x 10^9^/L, 21-50 x 10^9^/L, 51-100 x 10^9^/L and over 100 x 10^9^/L was calculated. **C.** Correlation between platelet A10 and platelet counts in ITP patients with (black circles) or without (open circles) symptoms of bleeding or bruising having a platelet count below 50 x 10^9^/L was calculated. Only significant p-value (p<0.05) highlighted in red are displayed. HDs (n=2-28). The primary ITP patient cohort (n=2–81) includes repeat measurements from the same subjects collected at different time points. Control group (n=11-20) includes patients with platelet count below 100 x 10^9^/L due to causes other than ITP. Two-way ANOVA with Bonferroni’s multiple comparisons test was performed and Pearson correlation coefficient (r) was calculated.

Platelet A10 correlated strongly with platelet count (r=0.797, p<0.0001, Figure 5B) and was significantly lower in symptomatic patients (p<0.0001, Figure 3H), those newly diagnosed (p=0.0135, Figure S3H), or those receiving first-line therapy (p=0.0005, Figure 4H).

To determine whether the platelet contribution to the clot was compromised in pITP, patients with bleeding and bruising symptoms were compared to those with equivalent counts and no symptoms. Correlation curves were constructed of the platelet A10 vs platelet count in patients with platelet counts below 50 x 10^9^/L. Moderate correlation with clinical symptoms was observed (r=0.727, p<0.0001, Figure 5C). Using OLS linear regression, the symptomatic patients were found to have a significantly lower platelet A10 level relative to the platelet count than the non-symptomatic patients (p=0.0027).

### Multivariable analysis

Given the multifactorial nature of bleeding in ITP, a multivariate approach was used to identify key predictors and account for confounding among clinical and laboratory variables. The goal was to determine whether a combination of biomarkers could better predict bleeding risk than platelet count alone. Bleeding and bruising symptoms in ITP were modeled using logistic regression, incorporating significant variables (p<0.05) in univariate analysis. To address multicollinearity and missing data, PPCA derived a composite score (PC1), explaining 42.6% variance (Table S4) and loading most strongly on platelet A10, TPO, GPVI, TLT-1, and CitH3-DNA (Table S5). Two logistic regression models were tested to predict the presence of symptoms in pITP (Table 2). In model 1, platelet count was used as the sole predictor of symptoms (p<0.01, β=−0.98) but demonstrated limited sensitivity (53.5%) and accuracy (70.5). Incorporating PC1 in model 2, improved model stability, accuracy (78.1% vs. 70.5%), sensitivity (69.8% vs. 53.5%) and AUC (0.87 vs. 0.83). PC1 emerged as the sole significant predictor (p=0.002), while platelet count lost significance (p=0.165), indicating a better overall fit (Figure S8).

**Table 2:** Multivariate logistic regression model and performance metrics.

| Predictor | Model 1 (Platelet count only) | Model 2 (PC1 included) |
| --- | --- | --- |
| Intercept (SE, P-value) | 2.433 (0.611, p<0.001) | 0.701 (0.783, p<0.001) |
| Platelet Count (SE, P-value) | -0.983 (0.201, p<0.001) | -0.367 (0.264, p=0.165) |
| PC1 (SE, P-value) | - | 1.194 (0.393, p=0.002) |
| Sensitivity | 0.535 | 0.698 |
| Specificity | 0.823 | 0.839 |
| Positive predictive value | 0.677 | 0.750 |
| Negative predictive value | 0.718 | 0.800 |
| Accuracy % (95% CI) | 70.48 (60.8-79.0) | 78.1% (69-85.6) |
| Kappa | 0.370 | 0.542 |
| Area under curve (AUC) | 0.83 | 0.87 |

PC1=principal component 1, CI=confidence interval, SE= standard error.

## Discussion

Patients with thrombocytopenia may experience bleeding events out of step with the platelet count for reasons which remain unclear, while other patients with severe thrombocytopenia may only experience minimal bleeding.^22,23^ Current diagnostic and treatment approaches focus on thrombocytopenia rather than investigating and addressing platelet dysfunction, primarily due to lack of reliable diagnostic tools to evaluate the pathology underlying thrombocytopenia. ITP is one such disease where, despite recent advances, platelet counts remain the principal determinant of clinical management with no definitive risk factors, and diagnostic or prognostic criteria.^23–25^ Here, we added a battery of research-based approaches to explore changes in platelet activation, function and other indices and found increased surface GPVI and TLT-1, increased serum TPO and plasma sGPVI and evidence of increased NETosis in bleeding patients with pITP. Even allowing for platelet counts, there were also differences in the platelet contribution to clot formation in bleeding patients implying a platelet dysfunction in pITP.

Subtle losses of receptor surface density can impact platelet activation and could underpin a mild bleeding propensity. Further, GPVI, TLT-1 and GPIbα can be metalloproteolysed^26^ and were reduced in a heterogeneous ITP cohort.^27^ These proteins were selected, as we recorded an increased expression of GPVI and TLT-1 in symptomatic patients that possibly suggest that their platelets were pro-adhesive and pro-aggregatory and contributed more to hemostasis compared to their asymptomatic counterparts. Although increased resting P-selectin expression in ITP platelets is reported,^23,28^ we did not observe a significant difference between patients with and without symptoms. This discrepancy may reflect differences in cohort characteristics, bleeding assessment methods, or technical aspects of P-selectin measurement.

Although newly-synthesised platelets were not elevated in the pITP cohort, a statistically significant positive relationship between GPIbα and the TO^bright^ population was found, consistent with links between GPIbα and circulating platelet age.^21^ This association likely reflects the preservation of GPIbα on younger, newly released platelets, which are less affected by receptor shedding or surface modulation that occur as platelets age, activate or with immune-mediated damage.

Exposure of new surface receptors may compensate for platelet loss and reduced haemostatic capacity in ITP. However, OG-Fg binding to activated platelets was stable across the subgroups. We used low and high doses of CRP-xL to assess subtle changes in GPVI-induced signalling and PAR-1 agonist and ADP were also used. pITP patients showed normal fibrinogen binding in response to all stimuli, indicating intact pathways regulating integrin αIIbβ3 activation.

This finding contrasts with previous reports that have described impaired integrin activation in ITP platelets and an association with bleeding risk,^28,29^ possibly explained by differences in assay methodology, patient selection, disease heterogeneity, and sample size. Treatment status and underlying mechanisms of thrombocytopenia may contribute to divergent findings. Our results underscore the complexity of platelet dysfunction in ITP and highlight the need for further studies using standardised methods across diverse cohorts to clarify the relationship between integrin activation and bleeding risk in ITP.

Changes in platelet receptor levels through proteolysis are associated with vascular injury^30^ and autoimmune disorders^31–33^ contributing to thrombocytopenia, bleeding and the generation of bio-specific markers. Here plasma sGPVI and sTLT-1 were measured. Elevated sGPVI in asymptomatic pITP patients compared to those with symptoms remains unexplained, but significant changes in patients on second-line and subsequent therapies may aid treatment stratification. No variation in sTLT-1 was observed by symptoms, disease duration, or treatment.

TPO levels in pITP were increased compared to HDs, aligning with other reported ranges.^34^ There was no variation in TPO levels based on the chronicity of the disease. However, elevated TPO levels were observed in patients who were symptomatic, had lower platelet counts, and were receiving first-line therapies for ITP. Furthermore, TPO levels in patients receiving TPO-RA were unchanged. These drugs share no sequence homology with native TPO^35^ and are not detected in the TPO ELISA.

Recent studies have underscored the fundamental role of NETs in systemic autoimmune disorders.^36^ We found elevated NET complexes in samples from symptomatic pITP patients. This may indicate an accelerated autoimmune response in these patients, amplifying NET formation to promote further autoantigen generation, possibly fueling ITP relapses, and observed bleeding in our cohort. However, whether the autoimmune response triggers NET formation, or NETs formed via activated platelets, initiate and perpetuate the immune response, remains to be addressed.^37^ Most studies focus on NETosis in prothrombotic states observed in ITP,^38,39^ NETs contribution to bleeding manifestations remains largely unexplored.

ROTEM offers insight into platelet and coagulation abnormalities with minimal sample processing.^40,41^ As anticipated, ROTEM parameters that are sensitive to platelet numbers (CFT, MCF, A10, and α-angle)^42^ were impaired in thrombocytopenic pITP samples. Correlations of bleeding score with clot firmness parameters in thromboelastography have been reported in adult and paediatric ITP.^43,44^ Using a ROTEM parameters *versus* platelet count standard curve, we assessed differences in platelet function between samples that had equivalent, low platelet numbers. Differential platelet A10 values in pITP samples with similar platelet counts were linked to bleeding incidence. These findings identify both a loss of ITP platelet haemostatic capacity and the utility of viscoelastometry to detect platelet dysfunction associated with bleeding in severe thrombocytopenic patients. However, analysis of ROTEM parameters in relation to other functional readouts revealed no significant correlations, suggesting that not all aspects of platelet dysfunction were captured by ROTEM alone. Platelet-bound antibodies were also evaluated in relation to functional parameters and activation markers (data not shown), but no significant differences were observed based on the presence or absence of these antibodies.

We used PPCA to identify underlying data patterns and develop predictive models. PC1 incorporation improved model accuracy and increased sensitivity by reducing missed symptomatic cases. PC1 (comprising largely of platelet and vascular activation markers) functions as a composite score that captures complex platelet-related mechanisms contributing to bleeding risk. Next steps will assess whether the model remains stable and generalisable across different patient populations. Further investigation into biological interpretations of PC1 may also provide insights into the underlying molecular mechanisms driving ITP bleeding risk.

Conducting a study with patients referred from a hospital setting presents certain limitations and selection biases. A meticulous selection of clinically annotated patients, acquisition of informed consent, transportation of samples to a research facility, contemporaneous collection of HDs, and periodic review of recorded diagnoses mandated a pragmatic approach. Most patient specimens were received on brief notice and expeditiously processed to conduct a wide range of platelet experiments on the day of collection. This constituted a formidable challenge to overcome and increased the validity of the findings. Although we analysed platelet function across different treatment modalities, no significant differences were observed; nevertheless, treatment heterogeneity remains a potential confounding factor that could influence platelet function and should be considered in the interpretation of the findings and in future studies.

In conclusion, elevated surface GPVI and TLT-1, serum TPO, plasma sGPVI, and NETs, and intriguing differences in platelet contribution to clot formation in symptomatic patients support the presence of platelet dysfunction in pITP. Multivariable analysis showed that a composite measure of platelet parameters predicted bleeding risk more accurately than platelet count alone. These findings underscore the complexity of platelet dysfunction in ITP, where no single biomarker reliably predicts bleeding. By leveraging multivariable models and composite scores,^45^ the interplay between platelet function, activation, and immune dysregulation using readily assayable vascular measures can be captured. These insights may improve risk stratification and guide personalised treatment, particularly as care shifts toward algorithm-driven, targeted approaches. Further validation with independent datasets and prospective studies is needed to confirm clinical utility.

## Data Availability

All data produced in the present study are available upon reasonable request to the authors.

## Acknowledgements

This work was supported by the National Health and Medical Research Council of Australia, the National Blood Authority of Australia, and ACT Health. SAA and SMH were recipients of scholarships from the Australian Government Research Training Program, and SAA received an ISTH Reach the World fellowship. The authors acknowledge Mrs Samina Nazir and Ms. Anila Jahangiri for their assistance with data collection, Dr. Harpreet Vohra and Mr. Michael Devoy from the JCSMR Cytometry, Histology, and Spatial Multiomics (CHASM) Facility for their support with data acquisition, the Canberra Health Services staff (bleeding disorders nurses Ms. Jayne Treagust, Ms. Lauren O’Connell, and Mr. James Slade, and hematology staff A/Prof. Philip Crispin and Dr. Jun Ng) for their contributions to patient sample collection and Prof. Steve Frost (University of Wollongong, Australia), Dr Teresa Neeman and Dr. Jonathan Roco (Biological Data Science Institute, ANU) for their statistical guidance.

## Authorship and conflicts of interest statements

SAA and SMH performed experiments, reviewed and interpreted the data and drafted the initial manuscript, LAC conceived aspects of the study, performed experiments and reviewed and interpreted the data; SAB, VB and YLT contributed to experimental data; RKA, EEG and PYIC conceived the study, acquired funding, reviewed the manuscript and interpreted the data. All authors reviewed the manuscript and approved its contents. PYIC received honoraria, speaking fees and travel assistance from Sobi, Novartis and Amgen; advisory boards for Janssen, Sanofi and Sobi; and research grants from Janssen and Novartis. EEG received speaking fees from Sobi. The other authors have no conflicts of interest to disclose.

## Supplementary Materials and Methods

### NPRC patient cohort

Diagnoses made by treating physicians were subject to revision as new clinical information became available over time. Three patients were reclassified as having Evans syndrome and categorised as secondary ITP. Thirteen patients who were initially suspected of having ITP were later found to have alternative causes of thrombocytopenia, including underlying liver disease, drug-induced thrombocytopenia, inherited platelet disorders, or cancer-related causes, and were reassigned to the control group. This approach ensured that final group allocations reflected the most accurate clinical diagnoses and maintained the robustness of group comparisons. Clinical histories, including bleeding assessments, comorbidities, medication use (such as NSAIDs or other agents known to affect platelet function) and thrombotic events were performed by the treating clinical team at sample collection. Documented arterial or venous thrombosis was present in 6/70 (8.5%) patients with pITP and 2/11 (18.1%) patients with sITP. While the overall frequency of thrombosis aligned with previously reported rates in ITP populations^46^, correlation analysis between thrombotic events and platelet parameters could not be reliably performed due to the limited number of events. Similarly, the utility of bleeding scores for biomarker correlation was limited by the narrow range of bleeding severity in the cohort, with most patients exhibiting only minor bleeding (score of 1). Accordingly, bleeding was analysed as a binary outcome (presence *versus* absence), and logistic regression models were used to evaluate associations between biomarker panels and bleeding symptoms. Samples were collected at the time of referral. To minimise sample processing and platelet loss, most experiments were carried out using anticoagulated WB, following a protocol designed to maximise platelet evaluation, alongside HD samples collected on same day.

### Blood drawing

Healthy donor blood was drawn using a 19G winged infusion set into syringes containing 3.2% trisodium citrate (TSC), 5 mM ethylenediaminetetraacetic acid (EDTA), or without anticoagulants for serum isolation. Patient blood was drawn into vacutainers containing TSC or EDTA (pink and white top tubes). Whole blood (WB) cell counts were obtained using an automated hematological analyser (Cell-Dyn Emerald 22, Abbott Diagnostics, USA). Blood samples were processed as soon as possible after collection, typically within 1-4 h, to minimise artefactual platelet activation and preserve physiological function.

### Reagents

Collagen-related peptide (CRP) monomer was purchased (Auspep, Melbourne, Australia) and chemically cross-linked as described.^47^ Thiazole orange (TO), trisodium citrate (TSC), adenosine diphosphate (ADP), ethylenediaminetetraacetic acid (EDTA), bovine serum albumin (BSA) and TWEEN®-20 were from Sigma-Aldrich (Merck, Darmstadt, Germany). Oregon green-488 fibrinogen (OG-Fg) was from Thermo Fisher (Waltham, MA, USA), PAR-1 agonist TRAP-6 was from R&D Systems (Minneapolis, MN, USA) and ROTEM reagents were from Werfen (Sydney, Australia). All other reagents were of laboratory grade.

### Antibodies

Phycoerythrin (PE) conjugated anti-αIIb monoclonal antibody (mAb), clone MEM-06, PE conjugated anti-CD62P mAb, clone AK4, and fluorescein isothiocyanate (FITC) conjugated anti-human IgG polyclonal antibody were sourced from Abcam (Melbourne, Australia). Allophycocyanin (APC)-conjugated anti-ADAM10, clone 163003, PE-conjugated anti-CD9, clone 209306, and FITC-conjugated anti-Trem-like Transcript (TLT)-1 mAbs, clone 268420 were obtained from R&D Systems (Minneapolis, MN, USA). FITC-conjugated anti-GPIbα mAb, clone AK2, was from Invitrogen (Thermo Fisher Scientific, Waltham, MA, USA), APC-conjugated anti-α2 integrin subunit mAb, clone P1E6-C5, was from BioLegend (San Diego, CA, USA), and FITC-conjugated anti-human IgG polyclonal antibodies were from eBioscience (California, USA). Anti-GPVI mAbs, clones 1A12 and12C9 and FITC-conjugated clone 1G5, have been previously described.^48^

### Reticulated Platelets

TSC-anticoagulated WB was diluted to a platelet count of 5 x 10^7^/mL in 137 mM NaCl containing 2.7 mM KCl, 10 mM Na2HPO4, 1.8 mM KH2PO4, 2 mM EDTA, pH 7.4 (phosphate buffered saline (PBS)-EDTA). Blood samples with a platelet count below 5 x 10^7^/mL were not diluted. Methanol-dissolved Thiazole orange (TO) was diluted to 0.5 ug/mL in PBS-EDTA then mixed with blood samples to a final concentration of 50 ng/mL. Samples were incubated at RT for 1.5 h in the dark then mixed with PE-anti-αIIb (1:250) and further incubated for 30 min. 5000 platelet events were acquired on a FACSCalibur. A gate collected the top 10% of CD41- and TO-positive events (TO^bright^) in a HD sample processed in parallel with each patient; the same gate was then applied to patient samples. The geometric mean of TO fluorescence was determined within this gated population. Recent studies have questioned the specificity of TO for identifying reticulated platelets due to potential confounding by activated platelets, large platelets, or microparticles^49,50^ We ensured reliable assessment of reticulated platelets by using stringent gating to minimise non-specific signals and a healthy donor with each patient sample.

### Resting and activated platelet αIIbβ3 measurement using OG-Fg

80 µL of TSC-anticoagulated WB was treated with 0.5 or 5 μg/mL CRP-xL, 10 µM thrombin receptor-activating peptide (TRAP)-6, or 5 μM ADP in the presence of 15 µg/mL of OG-Fg at RT for 2 h, protected from light. The 2 h incubation was selected to synchronise the completion of multiple platelet assays run in parallel, allowing all tests to conclude at a similar time and enabling simultaneous sample acquisition. After incubation, reactions were stopped by the addition of 50 mM EDTA and diluted 1:10 with 10 mM Tris-HCl pH 7.4, containing 150 mM NaCl and 5 mM EDTA (TS-EDTA). EDTA was added only after the activation and binding steps to terminate the reaction and prevent further platelet activation or aggregation; calcium was present during the binding phase to allow physiological fibrinogen-αIIbβ3 interaction. Fibrinogen binding to assess integrin αIIbβ3 activation was measured by the extent of fluorescence activity of OG-Fg using flow cytometry. This method directly measures the functional ligand-binding capacity of αIIbβ3, providing a physiologically relevant assessment. Platelets were gated using forward and side scatter properties and levels of OG-Fg binding across 5000 platelet events were assessed using the FL-1 channel. Positive OG-Fg gate was defined as fluorescence activity greater than that of unstimulated healthy donor samples. The same gate was applied to all other samples.

### Evaluation of platelet surface proteins

WB was used, which is well-suited for patients with low platelet counts and minimises sample manipulation. TSC-anticoagulated WB was diluted 1:5 with TS-EDTA and incubated with fluorescently conjugated antibodies against platelet-specific proteins at RT for 30 min, protected from light. Samples were diluted 2.5-fold with TS-EDTA, and antibody binding to 5000 platelet events (see Supplementary Figure S1 for gating strategy) was quantified on a FACSCalibur.

### Soluble GPVI ELISA

The assay was optimised using monoclonal mouse anti-human GPVI capture and detection antibody. Briefly, a 96-well plate (MaxiSorp, NUNC Denmark) coated with 12C9^48^ mAb against human GPVI, overnight at 4°C, was washed with PBS containing 0.2% TWEEN®-20 (PBS-T) and blocked with 1% (w/v) BSA. Duplicate aliquots of double-spun platelet-free plasma (PPP) were added for 1 h at RT. The plate was again washed with PBS-T and incubated with HRP-conjugated anti-human GPVI mAb 1A12. Bound antibody was quantified using enhanced chemiluminescence in a plate reader (Tecan Infinite 200Pro, Switzerland) by adding SuperSignal ELISA substrate (Pierce, Rockford, IL, USA) for 1 min. Amounts of sGPVI were interpolated against a standard curve constructed by mixing increasing amounts of N-ethylmaleimide (NEM)-treated platelet-rich plasma (PRP) with GPVI-depleted plasma.

### Measurement of serum and other plasma proteins

Serum thrombopoietin (TPO) levels were measured by ELISA (R&D Systems DTP00B). sTLT-1 levels were measured in platelet-free plasma by ELISA (R&D Systems DY2394). NETs were quantified by ELISA in plasma isolated from WB collected in a cell-free DNA collection tube containing EDTA (Roche 07785674001) by measuring levels of CitH3-DNA complexes following published methods.^51^

### Rotational thromboelastometry (ROTEM) for the evaluation of clot formation

Viscoelastography analysis of WB clotting parameters was performed using a ROTEM delta device (Werfen, Sydney, Australia) according to manufacturer’s instructions. The NATEM (using CaCl_2_ only), EXTEM (using CaCl_2_ and tissue factor), INTEM (using CaCl_2_, phospholipid, and ellagic acid), and FibTEM (using tissue factor and cytochalasin D) tests were performed with TSC-anticoagulated WB within 4 h of phlebotomy.

### Statistical analysis

Normally distributed data were analysed using unpaired t-tests or one-way analysis of variance (ANOVA) with Bonferroni’s multiple comparisons correction. Normality of residuals was assessed using D’Agostino and Pearson, Anderson-Darling, and Shapiro-Wilk tests. Non-normally distributed data were analysed using the Mann-Whitney U test or Kruskal-Wallis test with Dunn’s post hoc multiple comparisons test. Box and whisker plots illustrated minimum, median, and maximum values, along with the first and third interquartile ranges. Associations between platelet count and platelet A10 level in pITP patients with and without bleeding, was assessed using ordinary least squares (OLS) regression. The interaction between bleeding status and the association between platelet count and platelet A10 level was tested by incorporating an interaction term. Variables with p<0.05 in univariate analyses underwent logistic regression (symptoms as outcome) and continuous variables were log-transformed. Probabilistic principal component analysis (PPCA)^52^ addressed multicollinearity, imputed missing data, and derived composite scores. Model performance was assessed via ROC analysis (caret package).^53^ Analyses used R (v4.3.1)^54^ and GraphPad Prism (v10.2.1). Statistical significance was set at α=0.05, and Pearson correlation coefficient (r) was used to assess the strength of linear associations.

## Supplementary Tables

**Table S1:** Primary ITP classification based on clinical presentation and treatment.

| Primary ITP subgroup* | Newly Diagnosed<br>n=33 | Persistent<br>n=8 | Chronic<br>n=69 |
| --- | --- | --- | --- |
| **Bleeding/bruising<sup>†</sup>** |  |  |  |
| Yes (%) | 19 (58) | 4 (50) | 21 (30) |
| No (%) | 14 (42) | 4 (50) | 48 (70) |
| **On treatment<sup>†</sup>** |  |  |  |
| Yes (%) | 20 (61) | 4 (50) | 40 (58) |
| No (%) | 13 (39) | 4 (50) | 29 (42) |
| **On first line therapy<sup>‡</sup>** |  |  |  |
| Yes (%) | 17 (52) | 3 (38) | 7 (10) |
| No (%) | 03 (9) | 1 (13) | 33 (48) |

\*110 encounters from primary ITP patients, involving 70 individual patients, with repeated sample collections (n=21) at different time points, including during remission.
<sup>†</sup>Treatment includes steroids, IVIg, TPO-RA, rituximab, mycophenolate mofetil, dapsone, cyclosporine, danazol, vincristine, azathioprine, oseltamivir, ivalbumab, efgartigimod and/or splenectomy at time of blood collection.
<sup>‡</sup> Primary ITP patients receiving steroids and/or IVIg at the time of blood collection were classified as undergoing first-line therapy, while the remaining patients receiving other treatments were categorised as receiving second- or subsequent lines of therapy.

**Table S2:** Causes of thrombocytopenia in secondary ITP and control group.

| Likely causes of secondary ITP | n=11 |
| --- | --- |
| Drug induced* | 2 |
| Systemic lupus erythematosus | 2 |
| Evans Syndrome | 2 |
| ChAdOx1 vaccine-induced | 1 |
| STAT3 gain of function | 1 |
| Sjögren's syndrome | 1 |
| Paraneoplastic | 1 |
| Inflammatory bowel disease | 1 |
| **Likely causes of thrombocytopenia in control group** | **n=22** |
| Cancer related<sup>†</sup> | 6 |
| Liver disease<sup>‡</sup> | 4 |
| RUNX1 associated | 2 |
| Drug induced<sup>§</sup> | 1 |
| Inherited thrombocytopenia<sup>#</sup> | 2 |
| Other<sup>^</sup> | 7 |

\*Includes alemtuzumab and a checkpoint inhibitor.
<sup>†</sup>Includes acute myeloid leukemia, breast cancer, myelodysplastic syndrome, suspected gastrointestinal malignancy, thymoma and leiomyosarcoma.
<sup>‡</sup>Includes fatty liver disease, cirrhosis, hemi-hepatectomy and non-alcoholic steatohepatitis.
<sup>§</sup>Includes ruxolitinib and piperacillin / tazobactam.
<sup>#</sup> Includes grey platelet syndrome and MYH9 platelet disorder.
<sup>^</sup>Includes aplastic anemia, nutritional deficiency, bacterial (LPS)-induced and gestational thrombocytopenia.

**Table S3:**
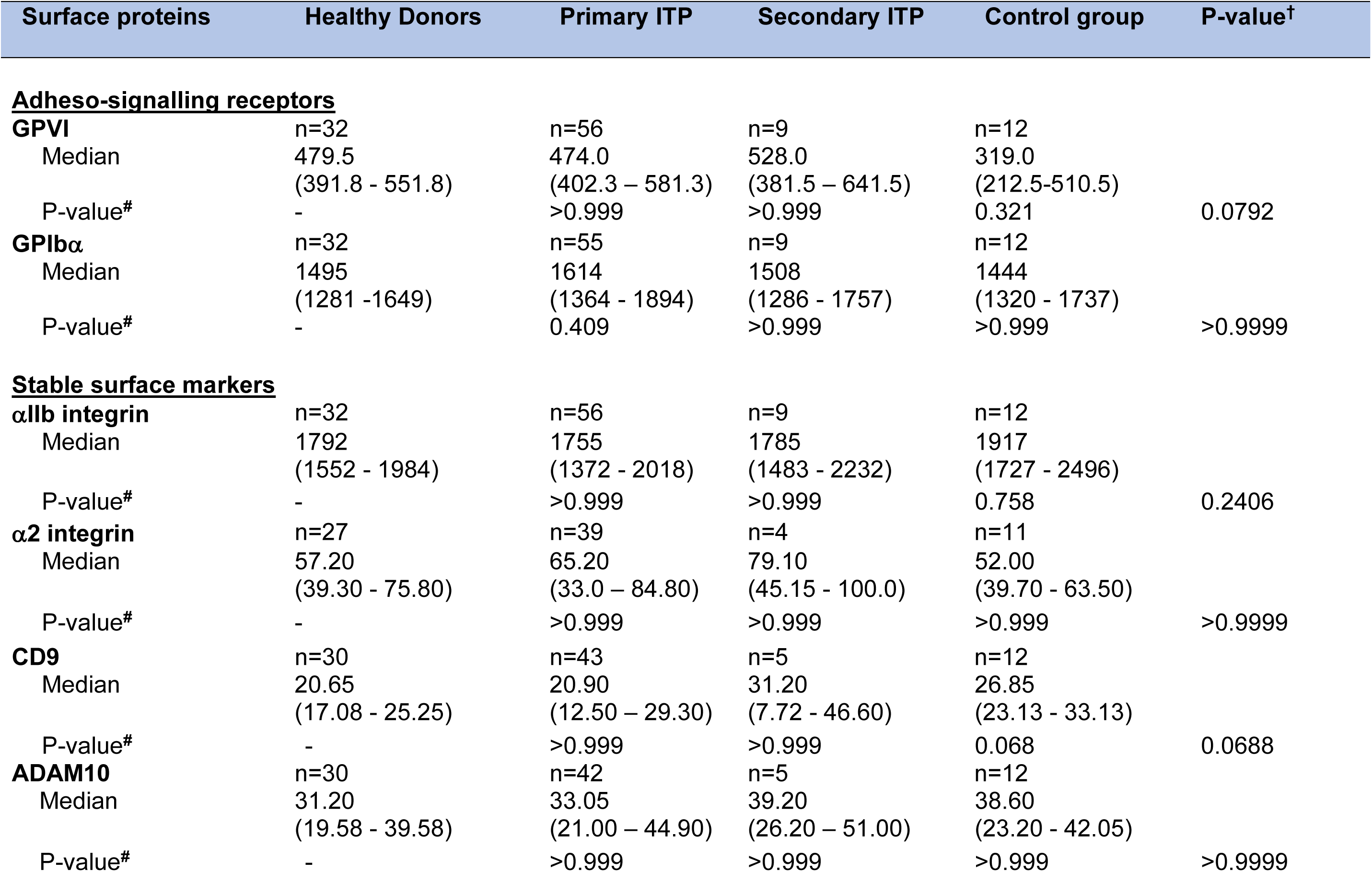
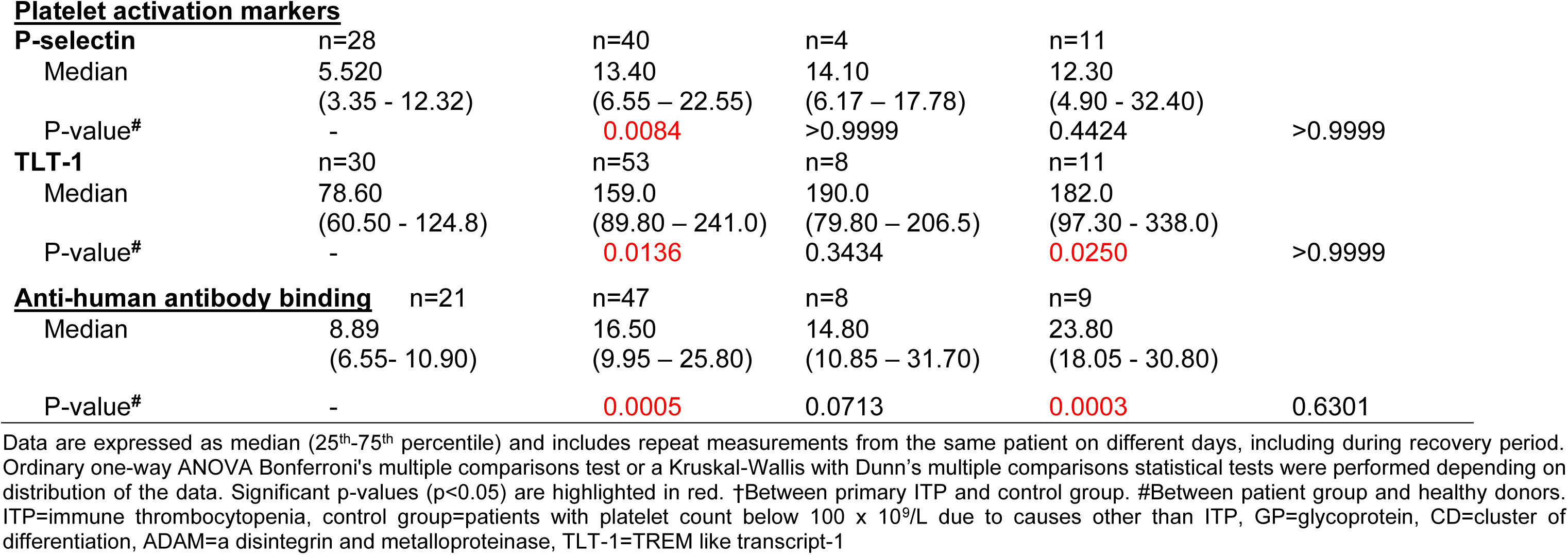
Platelet surface molecule expression in healthy donors, ITP and control group.

| Surface proteins | Healthy Donors | Primary ITP | Secondary ITP | Control group | P-value<sup>†</sup> |
| --- | --- | --- | --- | --- | --- |
| **<u>Adheso-signalling receptors</u>** |  |  |  |  |  |
| **GPVI** | n=32 | n=56 | n=9 | n=12 |  |
| Median | 479.5<br>(391.8 - 551.8) | 474.0<br>(402.3 – 581.3) | 528.0<br>(381.5 – 641.5) | 319.0<br>(212.5-510.5) |  |
| P-value<sup>#</sup> | - | >0.999 | >0.999 | 0.321 | 0.0792 |
| **GPIbα** | n=32 | n=55 | n=9 | n=12 |  |
| Median | 1495<br>(1281 -1649) | 1614<br>(1364 - 1894) | 1508<br>(1286 - 1757) | 1444<br>(1320 - 1737) |  |
| P-value<sup>#</sup> | - | 0.409 | >0.999 | >0.999 | >0.9999 |
| **<u>Stable surface markers</u>** |  |  |  |  |  |
| **αIIb integrin** | n=32 | n=56 | n=9 | n=12 |  |
| Median | 1792<br>(1552 - 1984) | 1755<br>(1372 - 2018) | 1785<br>(1483 - 2232) | 1917<br>(1727 - 2496) |  |
| P-value<sup>#</sup> | - | >0.999 | >0.999 | 0.758 | 0.2406 |
| **α2 integrin** | n=27 | n=39 | n=4 | n=11 |  |
| Median | 57.20<br>(39.30 - 75.80) | 65.20<br>(33.0 – 84.80) | 79.10<br>(45.15 - 100.0) | 52.00<br>(39.70 - 63.50) |  |
| P-value<sup>#</sup> | - | >0.999 | >0.999 | >0.999 | >0.9999 |
| **CD9** | n=30 | n=43 | n=5 | n=12 |  |
| Median | 20.65<br>(17.08 - 25.25) | 20.90<br>(12.50 – 29.30) | 31.20<br>(7.72 - 46.60) | 26.85<br>(23.13 - 33.13) |  |
| P-value<sup>#</sup> | - | >0.999 | >0.999 | 0.068 | 0.0688 |
| **ADAM10** | n=30 | n=42 | n=5 | n=12 |  |
| Median | 31.20<br>(19.58 - 39.58) | 33.05<br>(21.00 – 44.90) | 39.20<br>(26.20 – 51.00) | 38.60<br>(23.20 - 42.05) |  |
| P-value<sup>#</sup> | - | >0.999 | >0.999 | >0.999 | >0.9999 |
| **Platelet activation markers** |  |  |  |  |  |
| **P-selectin** | n=28 | n=40 | n=4 | n=11 |  |
| Median | 5.520<br>(3.35 - 12.32) | 13.40<br>(6.55 – 22.55) | 14.10<br>(6.17 – 17.78) | 12.30<br>(4.90 - 32.40) |  |
| P-value<sup>#</sup> | - | 0.0084 | >0.9999 | 0.4424 | >0.9999 |
| **TLT-1** | n=30 | n=53 | n=8 | n=11 |  |
| Median | 78.60<br>(60.50 - 124.8) | 159.0<br>(89.80 – 241.0) | 190.0<br>(79.80 – 206.5) | 182.0<br>(97.30 - 338.0) |  |
| P-value<sup>#</sup> | - | 0.0136 | 0.3434 | 0.0250 | >0.9999 |
| **<u>Anti-human antibody binding</u>** | n=21 | n=47 | n=8 | n=9 |  |
| Median | 8.89<br>(6.55- 10.90) | 16.50<br>(9.95 – 25.80) | 14.80<br>(10.85 – 31.70) | 23.80<br>(18.05 - 30.80) |  |
| P-value<sup>#</sup> | - | 0.0005 | 0.0713 | 0.0003 | 0.6301 |

Data are expressed as median (25<sup>th</sup>-75<sup>th</sup> percentile) and includes repeat measurements from the same patient on different days, including during recovery period. Ordinary one-way ANOVA Bonferroni's multiple comparisons test or a Kruskal-Wallis with Dunn's multiple comparisons statistical tests were performed depending on distribution of the data. Significant p-values (p<0.05) are highlighted in red. †Between primary ITP and control group. #Between patient group and healthy donors. ITP=immune thrombocytopenia, control group=patients with platelet count below 100 x 10<sup>9</sup>/L due to causes other than ITP, GP=glycoprotein, CD=cluster of differentiation, ADAM=a disintegrin and metalloproteinase, TLT-1=TREM like transcript-1

**Table S4:** Variance explained by principal components.

|  | PC1 | PC2 | PC3 |
| --- | --- | --- | --- |
| R<sup>2</sup> | 0.4257 | 0.2093 | 0.1406 |
| Cumulative R<sup>2</sup> | 0.4257 | 0.6349 | 0.7755 |

R<sup>2</sup>=coefficient of determination, PC=principal component

**Table S5:** PCA component loadings and composite scoring.

|  | PC1 | PC2 | PC3 |
| --- | --- | --- | --- |
| TPO | -0.541 | -0.334 | -0.183 |
| GPVI | -0.363 | 0.032 | -0.013 |
| TLT-1 | -0.306 | -0.011 | 0.019 |
| CitH3-DNA | -0.139 | -0.245 | 0.956 |
| sGPVI | 0.282 | -0.906 | -0.180 |
| Platelet A10 | 0.619 | 0.079 | 0.139 |

**Figure S1:**
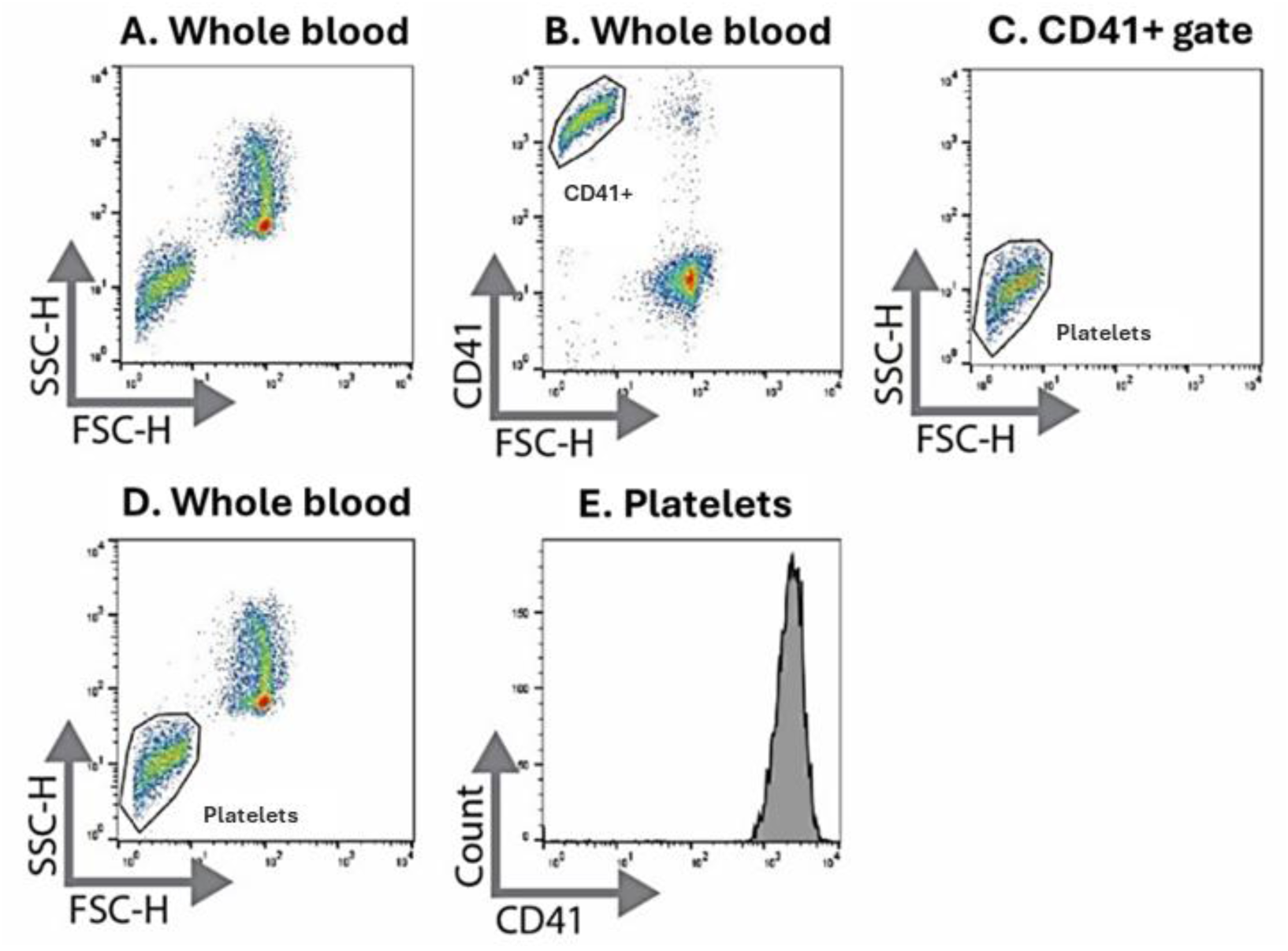
Gating strategy for platelet surface receptor enumeration. Flow cytometric data were acquired on a BD FACSCalibur. **A.** WB forward and side scatter are on x and y axes, respectively. **B.** The platelet gate is drawn around the CD41^+^ events only. **C.** Another gate is created around this population using forward and side scatter to be used for platelet events recorded with non-CD41 antibodies conjugated to various fluorophores. **D.** The new gate using forward and side scatter is applied to the WB. **E.** To validate that all events inside the platelet gate are CD41^+^, the platelet gate is assessed in a CD41^+^ histogram. The gating strategy was deemed satisfactory if at least 2000 events within the platelet gate were CD41^+^. FSC-H=forward scatter height, SSC-H=side scatter height, CD=cluster of differentiation.

**Figure S2:**
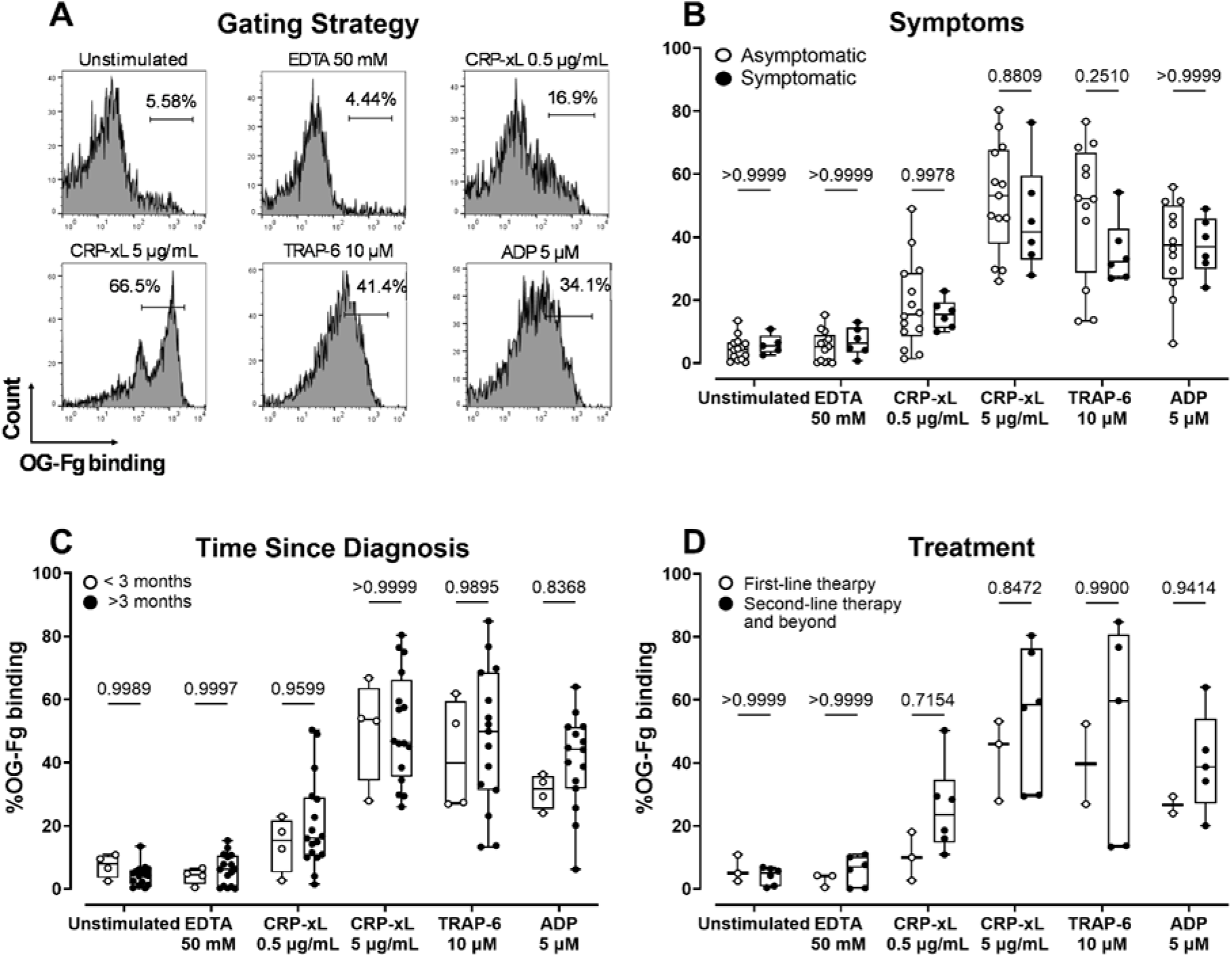
OG-Fg binding in primary ITP patient samples. OG-Fg binding was measured by flowcytometry (n=3-16). **A.** TSC-anticoagulated WB was activated 0.5 or 5 μg/mL CRP, 10 μM TRAP6 or 5 μM ADP and the binding of OG-Fg to activated platelets was measured by flow cytometry. Primary ITP patients are separated based on **B.** symptoms, **C.** time since diagnosis and **D.** first-line treatment (steroid and/or IVIg) at the time of sample collection. Data includes repeat measurement from the same subjects collected at different time points. A two-way ANOVA with Bonferroni’s multiple comparison test was performed.

**Figure S3:**
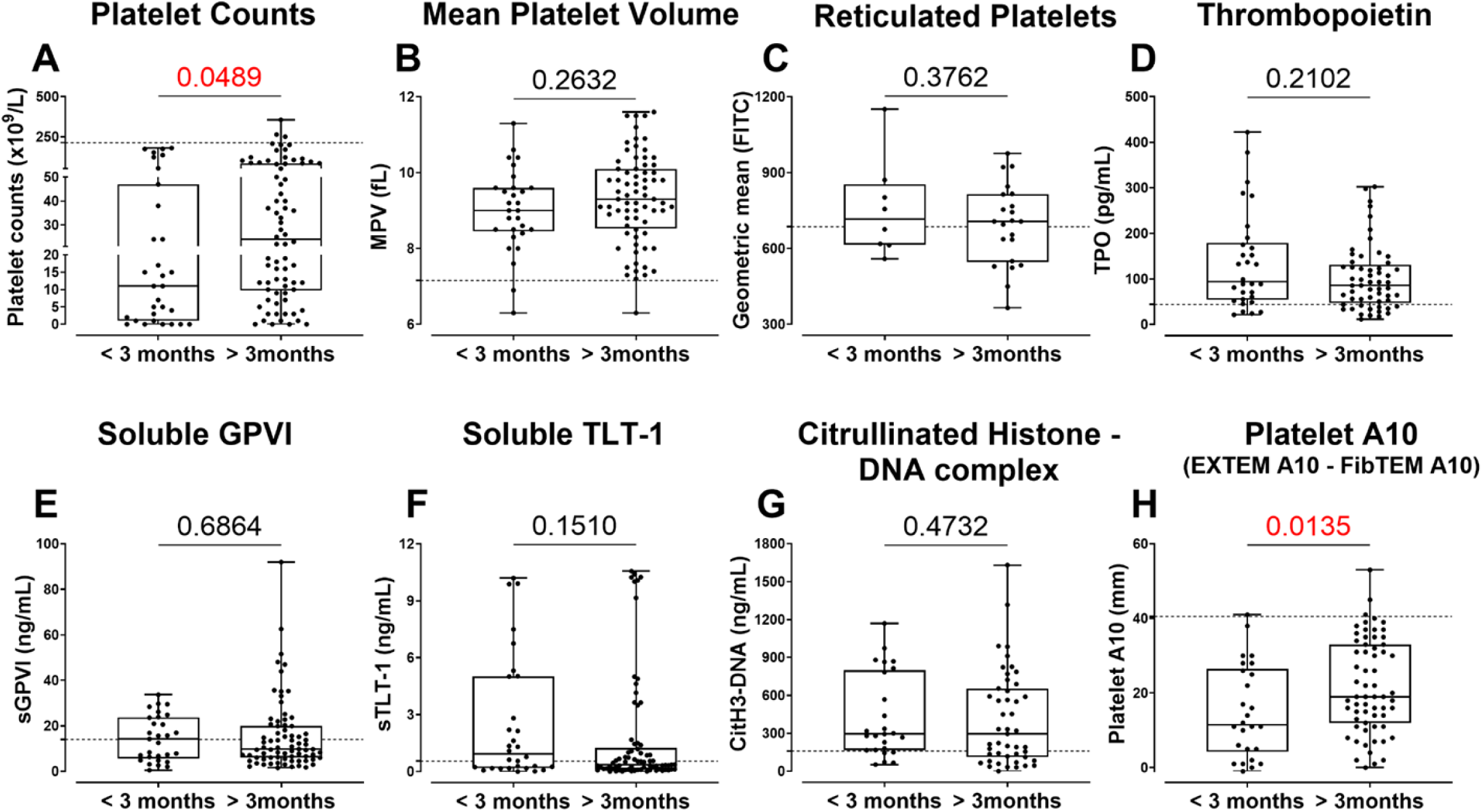
Platelet and plasma parameters in primary ITP patients stratified for duration since diagnosis. **A.** Platelet counts and **B.** MPV were measured using an automated haematology analyser in TSC-anticoagulated WB. **C.** Reticulated platelets were quantified by flow cytometry. Levels of **D.** TPO, **E.** sGPVI, **F.** sTLT-1 and **G.** CitH3-DNA complex were measured by ELISA. **H.** Platelet A10 was calculated by subtracting the FibTEM A10 measurement from the EXTEM A10 in ROTEM. The primary ITP patient cohort (n=8–75) includes repeat measurements from the same subjects collected at different time points. An unpaired t-test or Mann-Whitney test was performed depending on the distribution of the data. Significant p-values (p<0.05) are highlighted in red. The cutoff from HD levels is indicated by a dotted horizontal line on the respective graphs.

**Figure S4:**
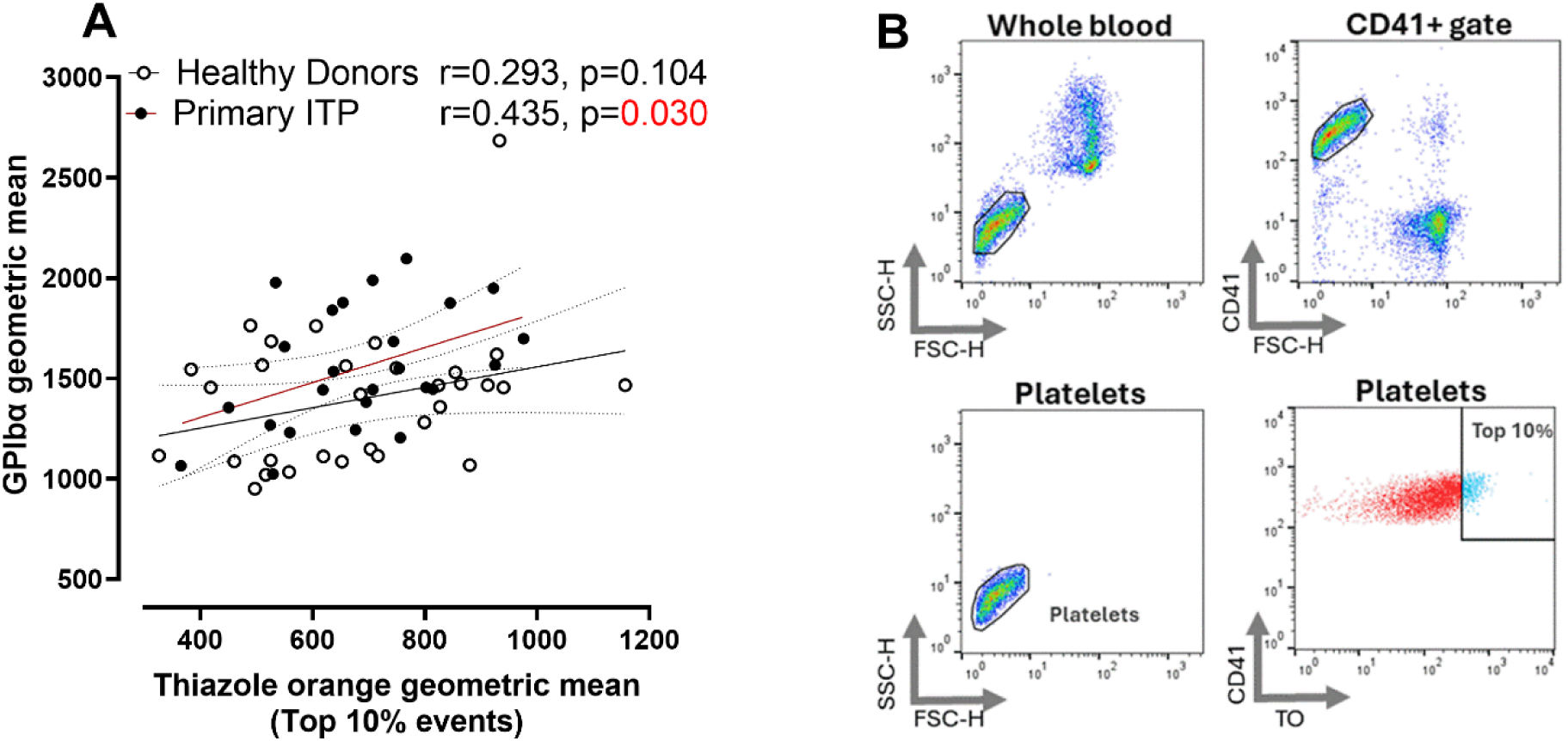
Correlation between reticulated platelets and surface GPIbα levels. Reticulated platelets and surface GPIbα levels were quantified by flow cytometry using thiazole orange (TO) and a fluorescently conjugated antibody respectively. **A.** The correlation between reticulated platelet and GPIbα levels was calculated for healthy donors and (n=32) and primary ITP patients (n=25). **B.** Gating strategy for TO staining of platelets. The top 10% TO^bright^ events were gated within CD41^+^ gate. Significant p-value (p<0.05) is highlighted in red. Simple linear regression was performed with 95% confidence interval (dotted lines) and Pearson correlation coefficient (r) was calculated. FSC-H=forward scatter height, SSC-H=side scatter height, CD=cluster of differentiation, TO=thiazole orange.

**Figure S5:**
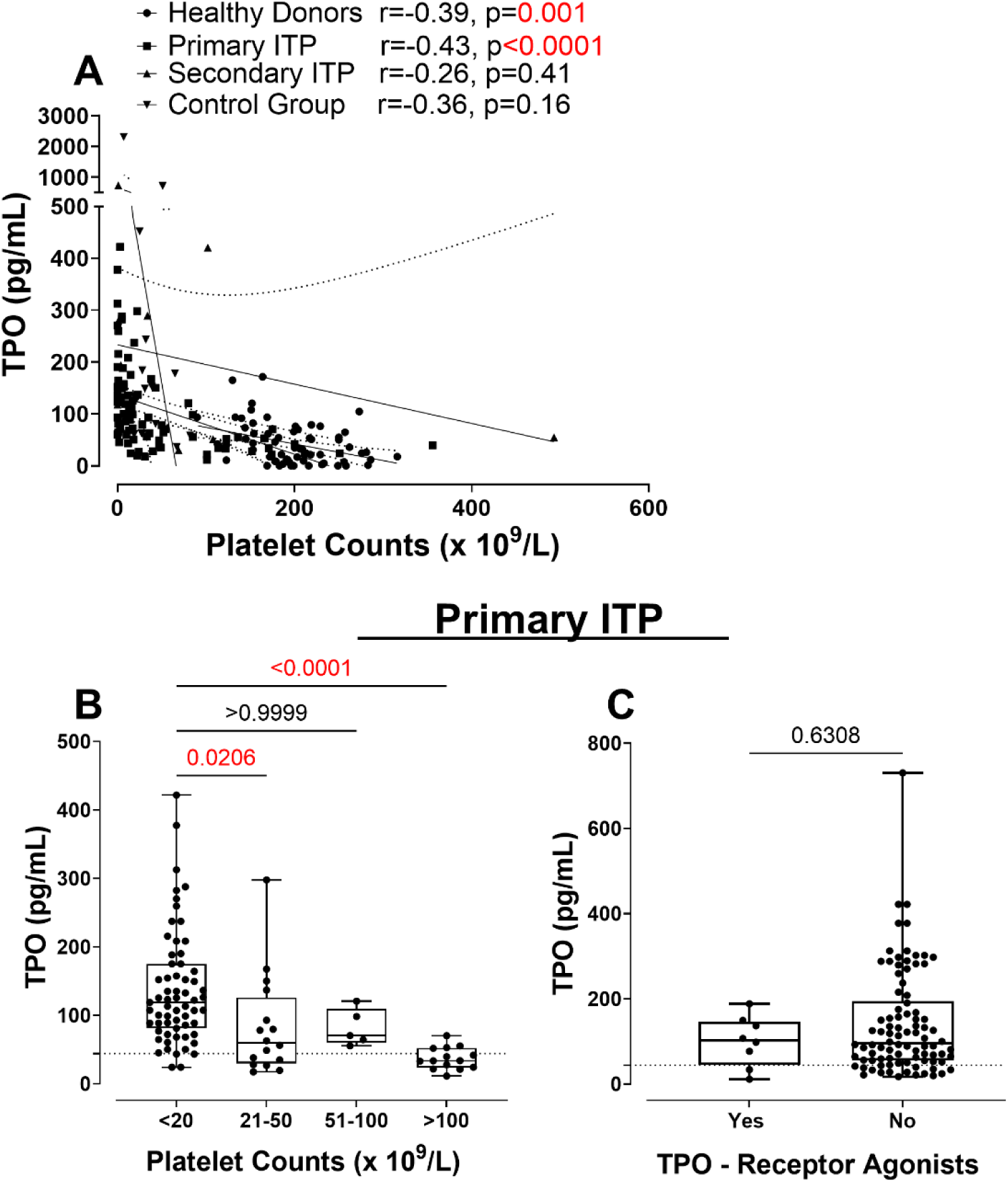
Thrombopoietin levels in healthy donors, ITP and control group. Serum TPO was measured by ELISA. **A.** Correlation of TPO with platelet counts was calculated. TPO concentrations in primary ITP patients were then categorised based on their **B.** platelet counts and **C.** TPO-receptor agonist intake at the time of sample collection. Data (n=5-88) includes repeat measurements from the same subjects collected at different time points. Kruskal-Wallis with Dunn’s multiple comparison test (A, B) and a Mann-Whitney test (C) were performed between groups. Significant p-values (p<0.05) are highlighted in red. The cutoff from healthy donor serum TPO levels, measured alongside these patients, is indicated by a dotted horizontal line on the respective graphs.

**Figure S6:**
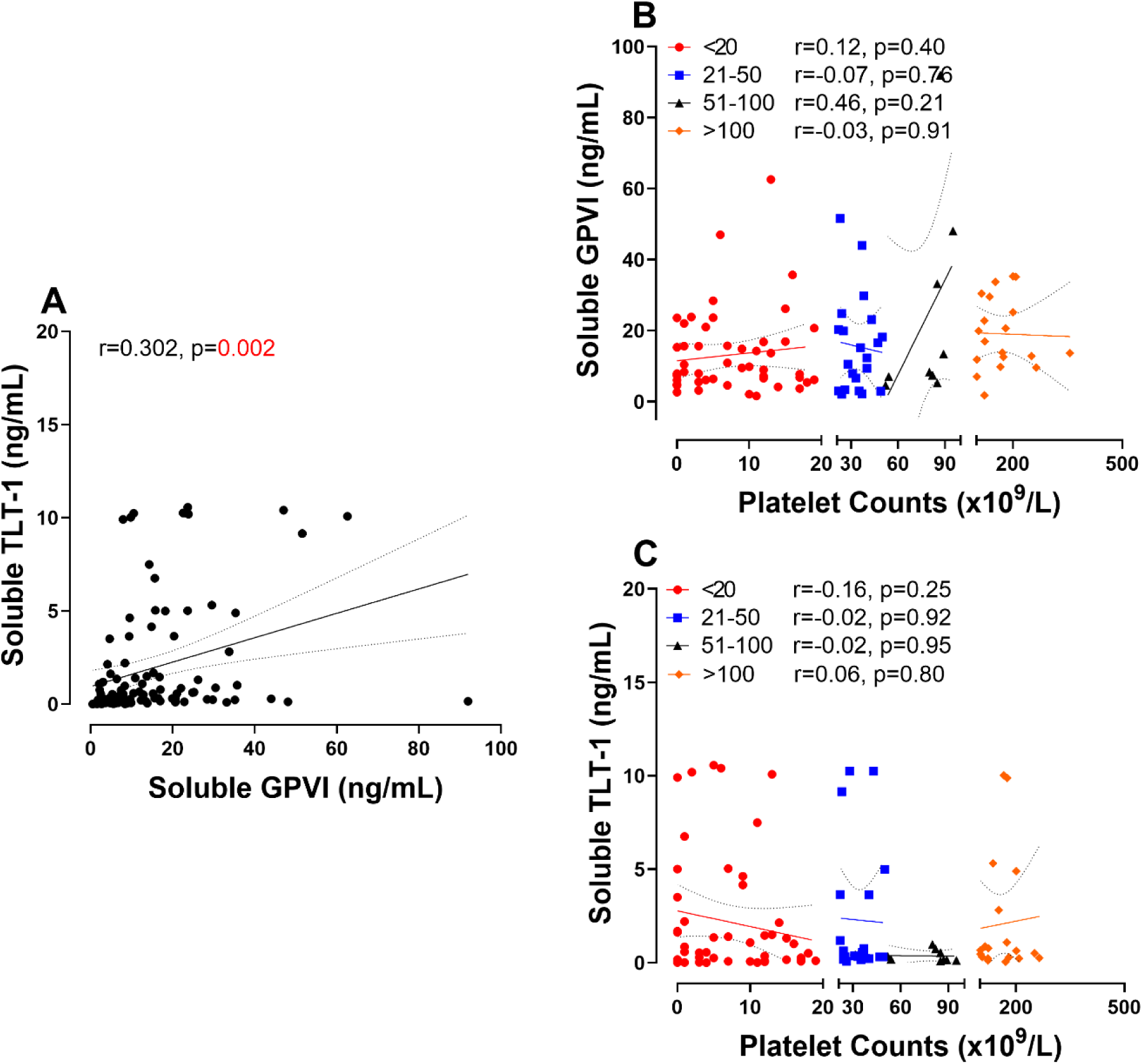
Correlation of soluble receptor levels with each other and platelet counts in primary ITP. Soluble TLT-1 and soluble GPVI were measured in platelet-free plasma (n=9-105). **A.** Correlation between soluble TLT-1 and soluble GPVI was calculated in primary ITP patients. Correlation between **B.** soluble GPVI and **C.** soluble TLT-1 in primary ITP patients with platelet counts less than 20 x 10^9^/L, 21-50 x 10^9^/L, 51-100 x 10^9^/L and more than 100 x 10^9^/L was calculated. Significant p-value (p<0.05) is highlighted in red. Simple linear regression was performed with 95% confidence interval (dotted lines) and Pearson correlation coefficient (r) was calculated.

**Figure S7:**
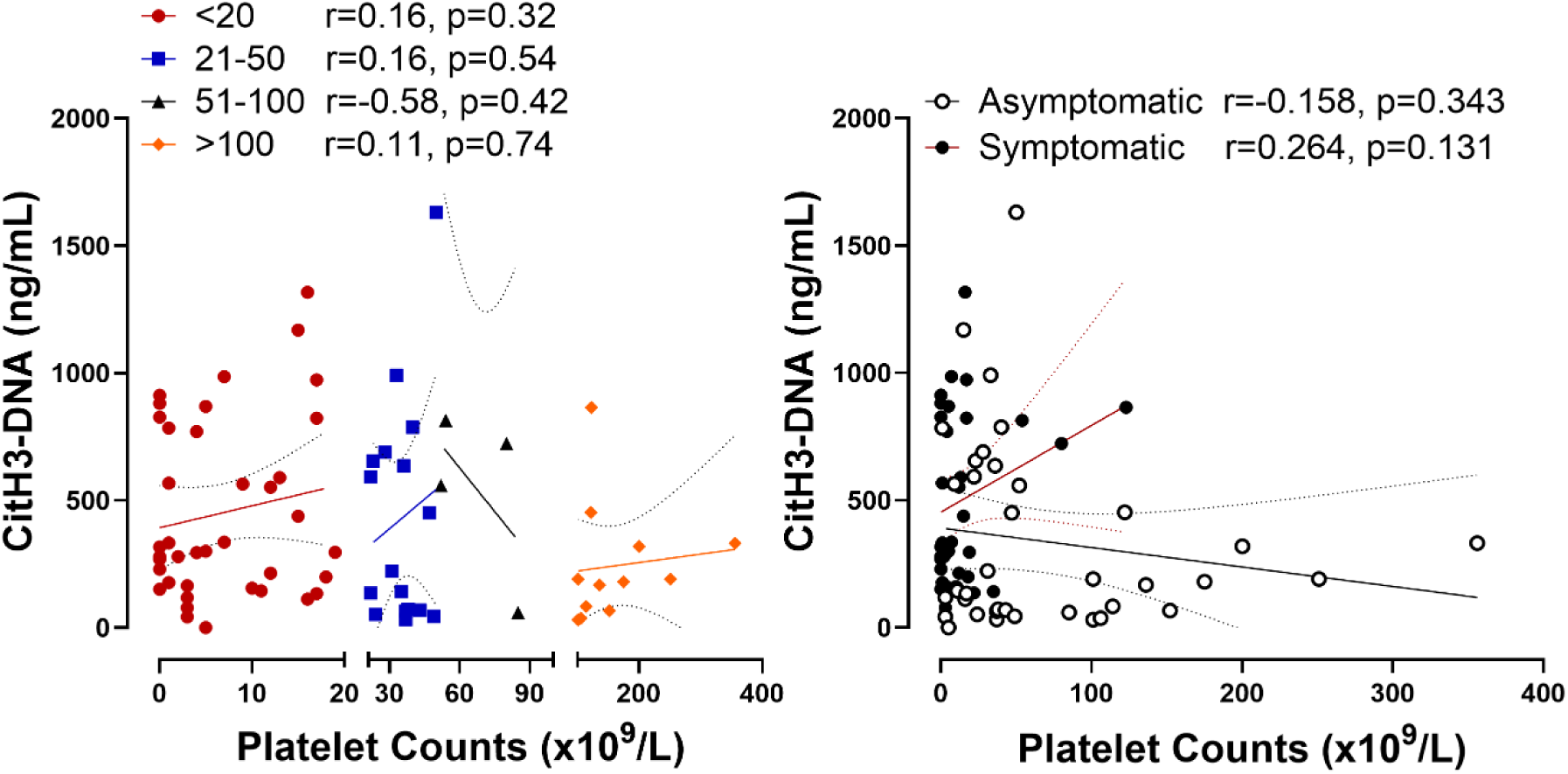
Correlation of CitH3-DNA complexes with platelet counts in primary ITP. CitH3-DNA complexes were measured in platelet-free plasma from whole blood collected in cell-free DNA collection tube (white top) with (n=4-39). **A.** Correlation between CitH3-DNA and platelet counts in primary ITP patients with platelet counts less than 20 x 10^9^/L, 21-50 x 10^9^/L, 51-100 x 10^9^/L and more than 100 x 10^9^/L was calculated. **B.** Correlation between CitH3-DNA and platelet counts in ITP patients with (black circles) or without (open circles) symptoms of bleeding or bruising. Simple linear regression was performed with 95% confidence interval (dotted lines) and Pearson correlation coefficient (r) was calculated.

**Figure S8.**
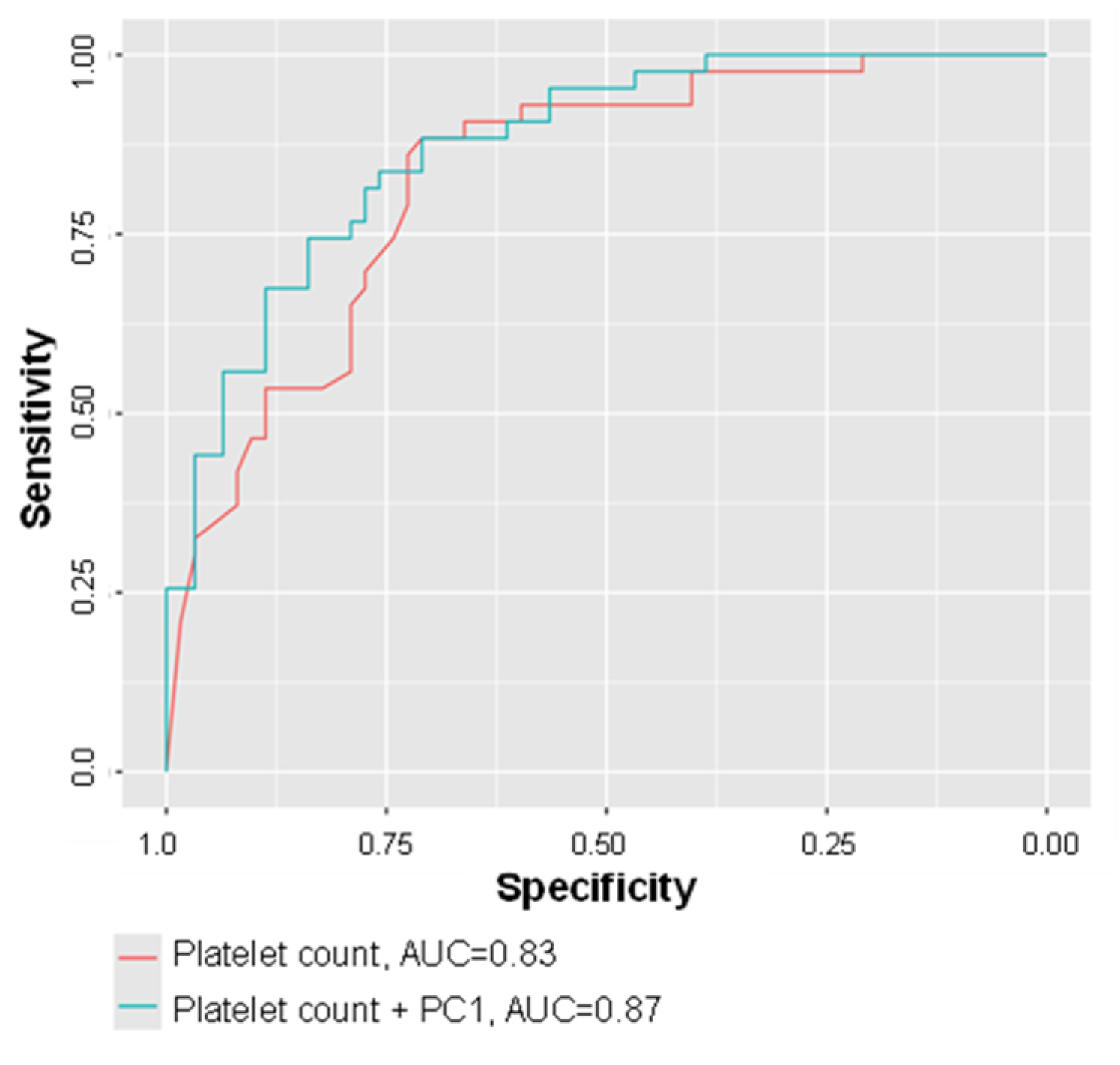
Receiver Operating Characteristic (ROC) curves for risk prediction models. The ROC curves compare the performance of two logistic regression models predicting the development of symptoms in primary ITP. Model 1 used platelet count as the sole predictor, while model 2 incorporated principal component 1 (PC1) alongside platelet count. The inclusion of PC1 improved model performance, reflected in a higher area under the curve (AUC), indicating enhanced predictive accuracy.

